# Cell-type-resolved chromatin accessibility in the human intestine identifies complex regulatory programs and clarifies genetic associations in Crohn’s disease

**DOI:** 10.1101/2024.12.10.24318718

**Authors:** Yu Zhao, Ran Zhou, Zepeng Mu, Peter Carbonetto, Xiaoyuan Zhong, Bingqing Xie, Kaixuan Luo, Candace M Cham, Jason Koval, Xin He, Andrew W. Dahl, Xuanyao Liu, Eugene B Chang, Anindita Basu, Sebastian Pott

## Abstract

Crohn’s disease (CD) is a complex inflammatory bowel disease resulting from an interplay of genetic, microbial, and environmental factors. Cell-type-specific contributions to CD etiology and genetic risk are incompletely understood. Here we built a comprehensive atlas of cell-type-resolved chromatin accessibility comprising 557,310 candidate cis-regulatory elements (cCREs) in terminal ileum and ascending colon from patients with active and inactive CD and healthy controls. Using this atlas, we identified cell-type-, anatomic location-, and context-specific cCREs and characterized the regulatory programs underlying inflammatory responses in the intestinal mucosa of CD patients. Genetic variants that disrupt binding motifs of cell-type-specific transcription factors significantly affected chromatin accessibility in specific mucosal cell types. We found that CD heritability is primarily enriched in immune cell types. However, using fine-mapped non-coding CD variants we identified 29 variants located within cCREs several of which were accessible in epithelial and stromal cells implicating cell types from additional lineages in mediating CD risk in some loci. Our atlas provides a comprehensive resource to study gene regulatory effects in CD and health, and highlights the cellular complexity underlying CD risk.

## Introduction

Crohn’s disease, one of the main types of inflammatory bowel diseases (IBD), is a chronic inflammatory disease of the gastrointestinal (GI) tract^1^. CD is highly heterogeneous in manifestation and can affect the entire GI tract, diminishing quality of life and decreasing life expectancy^2,3^. Incidence of CD is on the rise globally and is highest in the US and European countries^4^. While current treatments often achieve remission in patients, they do not cure CD, and inflammation recurs in most patients, often requiring surgical intervention^5^. Thus, there is an urgent need to better understand the cellular and genetic basis of CD and to develop effective strategies for therapeutic intervention and disease prevention.

While the etiology of CD is still poorly understood, it is generally believed to result from a complex interplay of host genetics with environmental factors and microbiota^6^. Untangling this interplay has been challenging due to extensive cellular heterogeneity within the gut and the contribution of multiple cell types to CD pathology. Adaptive and innate immune cells are linked to disease risk and severity^7^. Disruption of the epithelial barrier is characteristic of lesions in CD, directly implicating epithelial cell dysfunction in disease etiology^8,9^. Epithelial cells both react to pro-inflammatory signals and mediate inflammatory responses themselves, and cycles of epithelial healing and injury are characteristic of CD^10^. Fibroblasts are essential mediators in wound healing of the epithelium; however, in chronic inflammatory conditions they contribute to fibrosis and strictures^11,12^. Although CD is known to involve a wide range of cell types, delineating their individual contributions and disentangling causal roles from secondary involvement remains a major challenge in this complex disease.

Genome-wide association studies (GWAS) have linked more than 250 loci to IBD risk, providing a rich resource for untangling IBD biology and identifying therapeutic targets^13–16^. IBD risk loci and genes implicate a wide range of biological processes, including microbial sensing, adaptive immunity, and epithelial barrier function^9^. However, most of the putative causal variants are non-coding, thus making it challenging to identify their target genes or characterize their mechanism of action. Moreover, to translate these genetic signals into improved understanding of disease biology requires the identification of the precise cell types and contexts in which IBD-associated loci exert their effects on disease risk.

Single-cell RNA-seq (scRNA-seq) studies have used cell-type-resolved gene expression profiles to identify changes in cellular composition and gene expression associated with CD^17–19^, and to interpret genetic associations with IBD, revealing candidate genes and cell types that contribute to disease risk. Still, scRNA-seq alone is insufficient for prioritizing causal non-coding disease variants and nominating putative target genes based on gene expression level is complicated when genes are broadly expressed across cell types.

Open chromatin profiling – in particular, single-cell assay for transposase-accessible chromatin with sequencing (scATAC-seq) – can complement studies of single-cell gene expression by capturing active regulatory elements that are not directly profiled by scRNA-seq. Indeed, atlases of candidate cis-regulatory elements (cCREs) have led to the discovery of cell-type-specific regulatory features, linked complex disease risk to specific cell types and pathways^20–28^, and identified quantitative trait loci associated with chromatin accessibility (caQTLs)^29,30^. Importantly, recent cellular atlases of gut mucosa in healthy adults and children with CD, respectively, demonstrated that cell-type-resolved chromatin accessibility in the gut can provide some insights into cell types mediating IBD risk^31–33^.

Here we generated a cell-type-resolved atlas of chromatin accessibility of the intestinal mucosa using pinch biopsies obtained from the ascending colon (AC) and terminal ileum (TI) of individuals with and without CD (Fig. 1a). We used this atlas to identify and characterize gene regulatory programs in active CD (Fig. 1b), which allowed us to identify cell-types that mediate CD risk and prioritize non-coding CD risk variants (Fig. 1c).

**Fig. 1.**
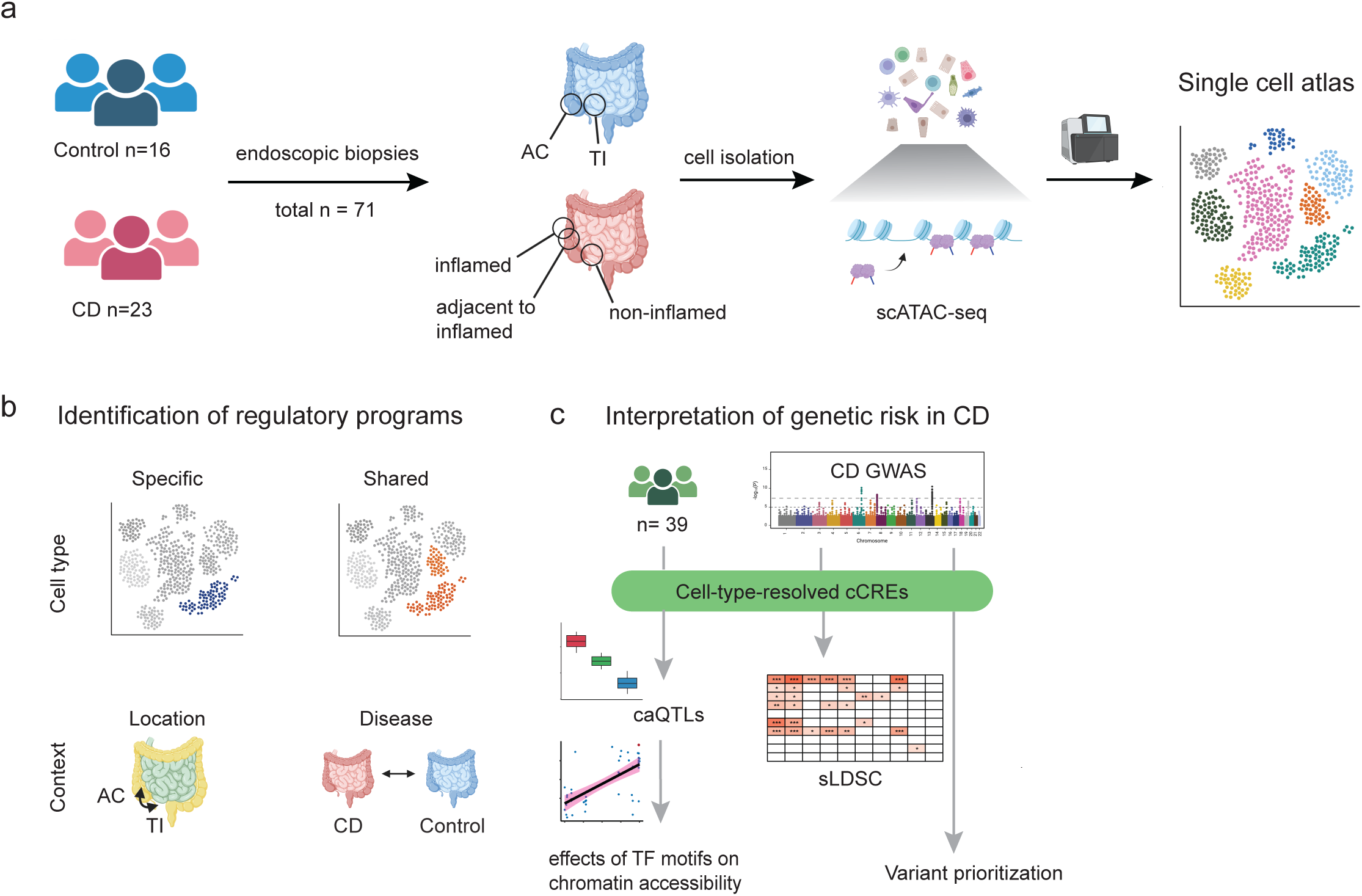
S**c**hematic **overview of study design. a**, A total of 71 biopsies were collected from the terminal ileum or ascending colon of 16 control individuals and 23 Crohn’s disease (CD) patients. These biopsies were stratified into four groups: control tissue, non-inflamed tissue from CD patients, non-inflamed tissue adjacent to inflamed regions, and inflamed tissue. Cell-type-resolved atlas of chromatin accessibility was constructed using scATAC-seq data from each biopsy. **b**, Schematic illustrating biological contrasts explored using this scATAC-seq atlas. **c**, Schematic outlining integrated and complementary approaches to understand effects of common genetic variants on chromatin accessibility, measure cell-type-specific IBD risk, and prioritization of risk variants and cell-types through integration of these data with finemapped CD GWAS variants.

## Results

### A regulatory atlas of the intestinal mucosa

We obtained terminal ileum (TI) or ascending colon (AC) biopsies from 23 CD patients and 16 healthy controls which we used to isolate single cells and perform scATAC-seq (Fig. 1a, Extended Data Fig. 1). After quality control we retained 178,030 cells and followed an established computational workflow to cluster the data using ArchR^34^ (**Methods**). We distinguished 29 clusters which we annotated with cell-type labels from matched scRNA-seq data (Methods, Fig. 2a, Extended Data Fig. 2); in-depth analysis of the scRNA-seq data is described elsewhere (Zhou et al.*, in preparation*). These cell type clusters show characteristic chromatin accessibility patterns at loci of previously published marker genes^18,35,36^ (Methods) (Fig. 2c).

**Fig. 2.**
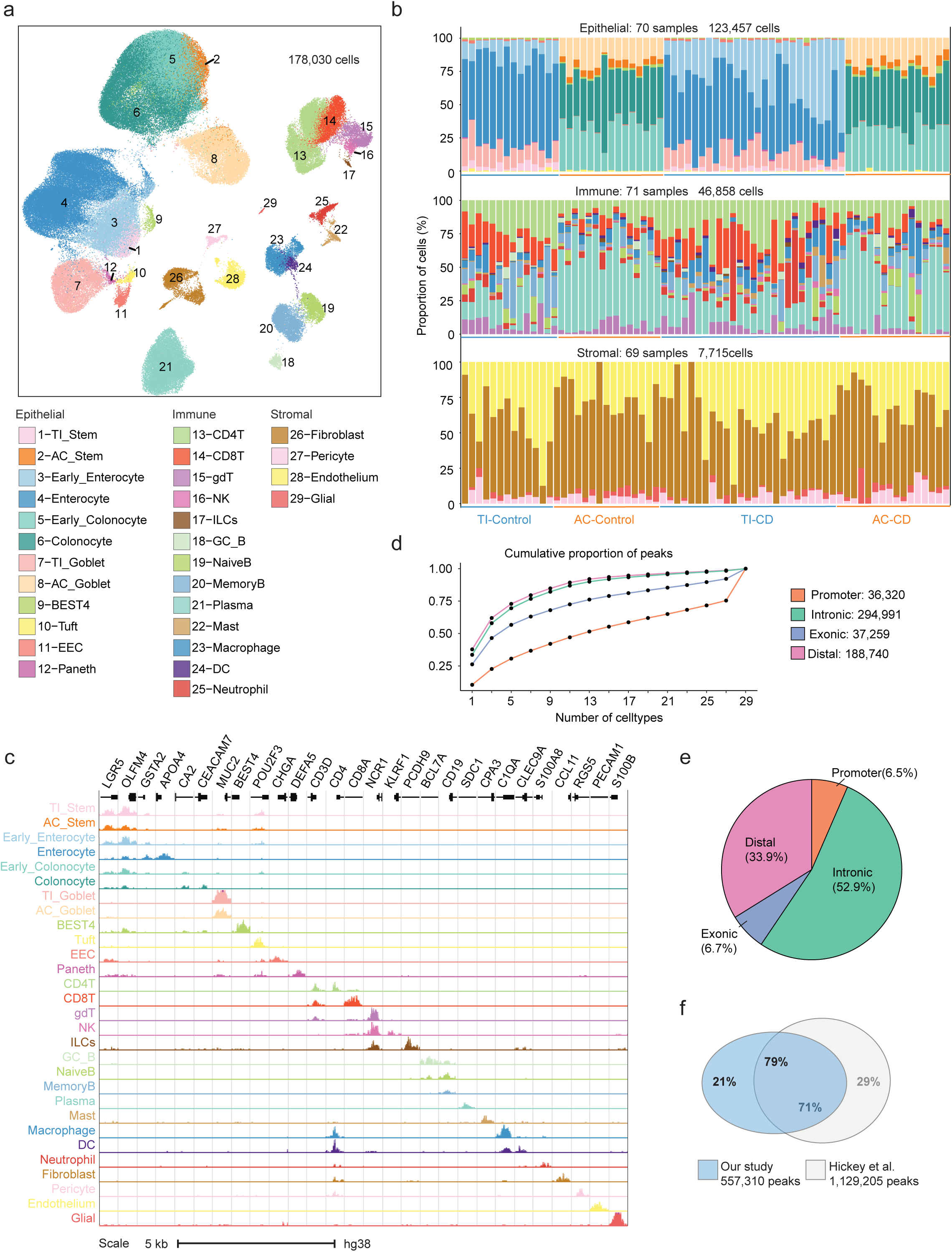
A**t**las **of cell-type-resolved chromatin accessibility in the human gut. a**, UMAP visualization of 178,030 cells from 29 distinct cell types across epithelial, immune and stromal lineages in the terminal ileum and ascending colon. **b**, Sample-wise proportions of cell type clusters separated by lineage (epithelial, immune, stromal). **c**, Normalized chromatin accessibility at differentially accessible regions (DARs) near marker genes for each cell type cluster. Gene names and structures are depicted above the accessibility tracks. Individual loci are separated by grey vertical lines. **d**, Comparison of the proportion of peaks shared between cell types in different peak categories (promoter, distal, intronic, exonic). Shown is the cumulative proportion of accessible peaks shared in a given number of cell types in each of the different categories. **e**, Proportions of peaks overlapping distinct genomic categories (promoter, distal, intronic, exonic). **f**, Proportion of overlap between the union peak set generated in this study and the union peak set published in Hickey et al^31^.

Our atlas captures cells of the epithelial, immune, and stromal lineages and represents all major cell types previously identified in transcriptome-based cellular atlases^31,35,37,38^. Epithelial cells represent the largest proportion of cells in our dataset (69.3%, 123,457 cells), followed by immune cells (26.3%, 46,858 cells) and stromal cells (4.3%, 7,715 cells) (Fig. 2b). Specifically, we identified 12 epithelial cell clusters, most of which further separate into distinct clusters based on their site of origin (AC and TI) (Fig. 2a, c, Extended Data Fig. 3a, b). These include large clusters of mature absorptive cells in the TI (Enterocytes) and AC (Colonocytes), as well as progenitor and stem cell populations. We also found prominent clusters of goblet cells from both locations. In addition, we detected clusters representing smaller cell populations, including Paneth cells which are restricted to the TI, enteroendocrine cells (EECs), and Tuft cells. In contrast to epithelial cells, immune and stromal cells do not separate based on site of origin (Extended Data Fig. 3 c, d, e, f).

Major cell types of adaptive immunity are CD4+ T cells and CD8+ T cells, ψ8T cells, NK and innate lymphoid cells (ILCs), plasma B, memory B, naive B, and germinal center B cells. Among innate immune cells we detected macrophages, mast, neutrophils and dendritic cells (DCs). Plasma B cells are most abundant (30%) among immune cells, followed by CD4+ and CD8+ T cells (Fig. 2b). We observed four distinct cell types within the stromal lineage, corresponding to fibroblasts, endothelial cells, pericytes, and glial cells. Due to the smaller number of stromal cells, we did not attempt to subcluster these further.

To identify cCREs for each cell type, we called peaks of accessible chromatin using MACS2^39^ on each cluster separately (Methods). We combined all peaks into a union set comprising 557,310 cCREs (Extended Data Fig. 4a). Most of these are located in introns (52.9%) or distal intergenic regions (33.9%), while 6.5% are located within or proximal to promoters (Fig. 2e). As observed previously, cCREs in or near promoters are shared across more cell type clusters compared to distal and intronic peaks^40^ (Fig. 2d). A large proportion of all identified cCREs (79%) overlap regions reported in a multiomic reference atlas of the healthy human intestine^31^ (Fig. 2f). To test which transcriptional regulators are associated with regulatory features in each cell type cluster, we performed motif enrichment analysis on cell-type-specific peak regions (Extended Data Fig. 4b) (Methods). Many of the top-ranking motifs in each cell type correspond to transcription factors (TFs) known to be master regulators in that cell type (e.g., HNF4a in enterocytes and RUNX3 in T cells), and motif accessibility is correlated with expression level of the respective TF across cell types (Extended Data Fig. 4c). This analysis confirmed that the cell-type-resolved cCREs capture sequence motifs of cell-type-specific transcriptional regulators. Together, these data represent a comprehensive atlas of cell-type-resolved cCREs in the human intestinal mucosa comprising samples from healthy adult and CD patients.

### Cell-type-specific, shared, and context-specific regulatory features in the intestinal mucosa

Discrete cell type clusters do not fully capture the complex and dynamic gene regulatory regions and programs in the intestinal mucosa which might be shared across cell types or associated with continuous processes (e.g., differentiation), contexts (e.g., inflammation) or location (e.g., AC vs TI). To complement the cluster-based cell type annotations we therefore adopted a grade of membership approach based on topic modeling^41–43^. By allowing for partial memberships, this approach is able to model continuous or indistinct boundaries between cell types, and by allowing for membership in multiple topics, it is able to model regulatory processes that might be shared across multiple cell types. To avoid over-representation of abundant cell types we downsampled each cluster to a maximum of 100 cells per individual (Methods). Examining the accessible regions associated with each topic, we leveraged enriched TF binding motifs to infer active TFs, and identify their putative target genes (Methods, Supplementary Table 3).

The majority of topics capture specific cell types or multiple closely related cell types within a particular lineage. In addition, we identified topics capturing shared effects, e.g., associated with biopsy location and inflammation status. Finally, several topics are shared broadly across cell types capturing basic features of the gene regulatory architecture.

We considered a topic to be cell type specific if the average topic proportion for cells within a given cluster was at least 60% and 5-fold higher than for the second highest cluster (Methods). Topics defined primarily by a cell type are typically enriched for well-known motifs and genes associated with that cell type. For example, Topic 5 is associated with Goblet cells in the TI (Fig. 3a, b, Extended Data Fig. 5a) and topic-peak regions are enriched for motifs of FOXA1/2/3 and KLF3/4/5. Putative target genes of this topic include *MUC2* and *FCGBP* and are enriched for O-linked glycosylation, maintenance of gastrointestinal epithelium, and other functions (Fig. 3b, c, Supplementary Table 3). Of note, we also detected many topics that are shared between related cell type clusters or associated with specific contexts in our dataset (Fig. 3a, b). For example, Topic 18 is strongly enriched in DCs and to a lesser degree, macrophages, likely reflecting the shared origin and overlapping function of these cell types. The associated accessible regions are enriched for motifs of TFs active in DCs, including SPI1 and IRF4, and are located near genes enriched for antigen presentation and chemokine signaling pathways (Fig. 3a, c, Extended Data Fig. 3a, Supplementary Table 3). Topic 29 captured regulatory features associated with both early and mature colonocytes (Fig. 3a, b). These regions are enriched for motifs associated with TFs active in colonocytes (e.g., CDX1/2) (Fig. 3a, Extended Data Fig. 3a, Supplementary Table 3) whose targets include genes known to be expressed in colonocytes (e.g., *MS4A12*, *CA2*, and *SATB2*; see Fig. 3c, Extended Data Fig. 3a, Supplementary Table 3).

**Fig. 3.**
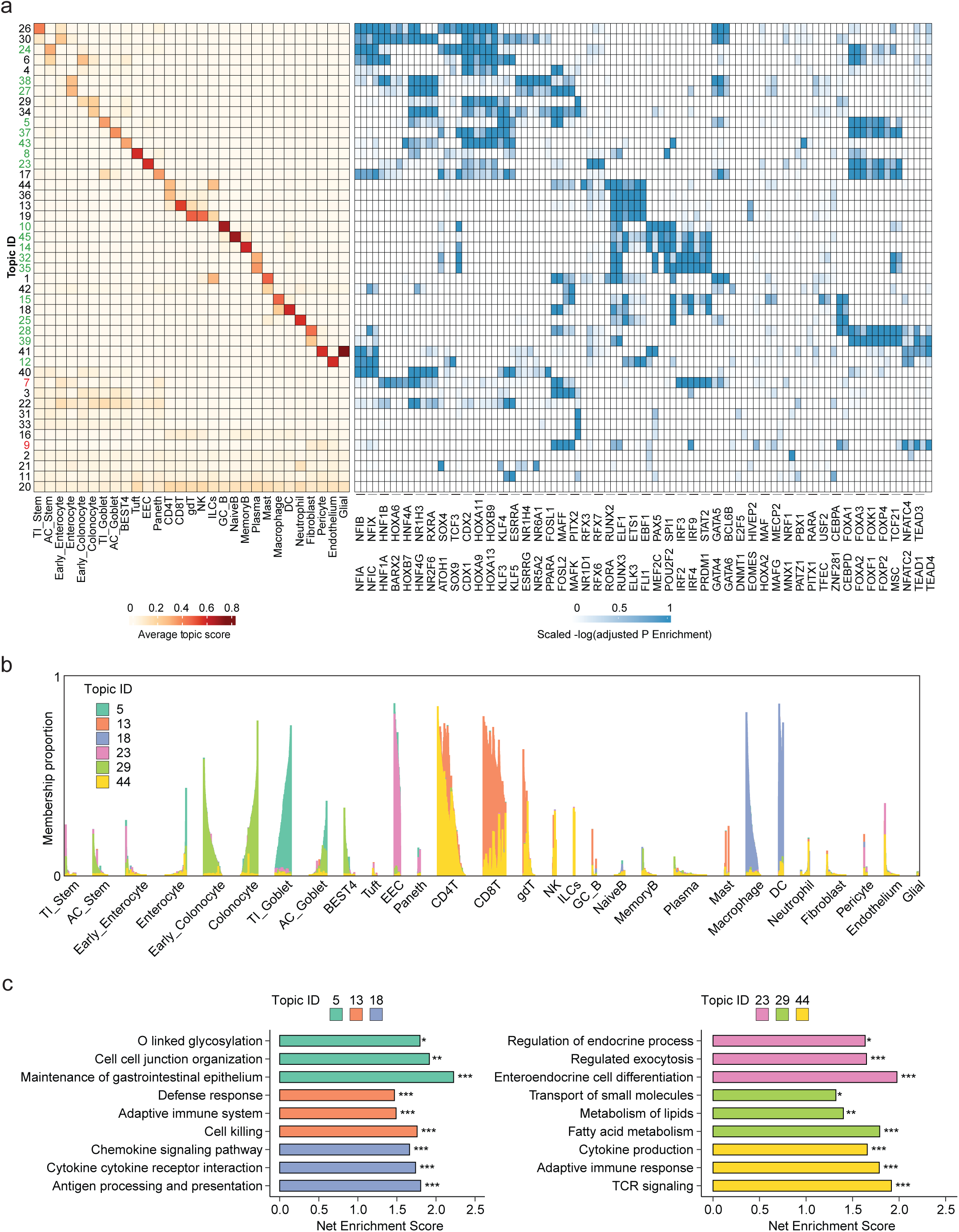
I**d**entification **of shared, cell-type-, and context-specific regulatory programs through topic modeling. a**, Average topic membership for cells grouped by cell type clusters (left). Topics specific to cell type and inflammation are labeled in green and red, respectively. TF motif enrichment for each topic (right). Shown are the significant enriched TF motifs for each topic (adjusted p-value < 0.01). The -log10(adjusted p-value) for motif enrichment was capped at a maximum of 30, with any values exceeding 30 being set to 30. These capped values were then scaled by the maximum value for each TF and plotted. **b**, Structure plot displaying proportion of selected topics in individual cells across all cell types. Each column represents a single cell and cells are grouped by cell types. **c**, Pathway enrichment analysis based on genes associated with each of the selected topics. Pathways significantly enriched for intestine-related biological functions (FDR < 0.05) were selected and visualized. (***: FDR ≤ 0.001, **: FDR ≤ 0.01, *: 0.01 < FDR ≤ 0.05)

Finally, topics 11 and 2 are shared across all cells and represented mostly cCREs with invariant chromatin accessibility across cell types, such as promoters (promoter cCREs make up 38% of topic 11-specific cCREs while the average proportion of promoter peaks is 9.2% (s. d. =11%) across all topics) and distal elements enriched for CTCF motifs (topic 2) (Fig. 3a, Extended Data Fig. 5a, Supplementary Table 3). Topic analysis therefore captures regulatory features that are also identified using discrete cell type clusters. In contrast to cluster-based analysis alone, however, we detected many topics that are shared between related cell type clusters or associated with important biological variables in the intestinal mucosa in healthy individuals and CD patients (Fig. 3a, b).

### Pervasive changes in regulatory landscape during inflammation in the epithelial lineages

Inflammation is a defining characteristic of CD and underlies its clinical symptoms. To discover cell-type-specific regulatory programs involved in mucosal inflammation, we leveraged topic analysis to identify inflammation-associated cell types, transcriptional regulators, and cCREs.

We found two topics, topics 7 and 9, to be strongly associated with inflammation. Topic 7 proportion is significantly increased in epithelial cells in biopsies taken directly from inflamed regions but shows only moderate enrichment in patient-matched biopsies from non-inflamed adjacent regions and none in biopsies from healthy controls (Fig. 3a, 4a, b).

**Fig. 4.**
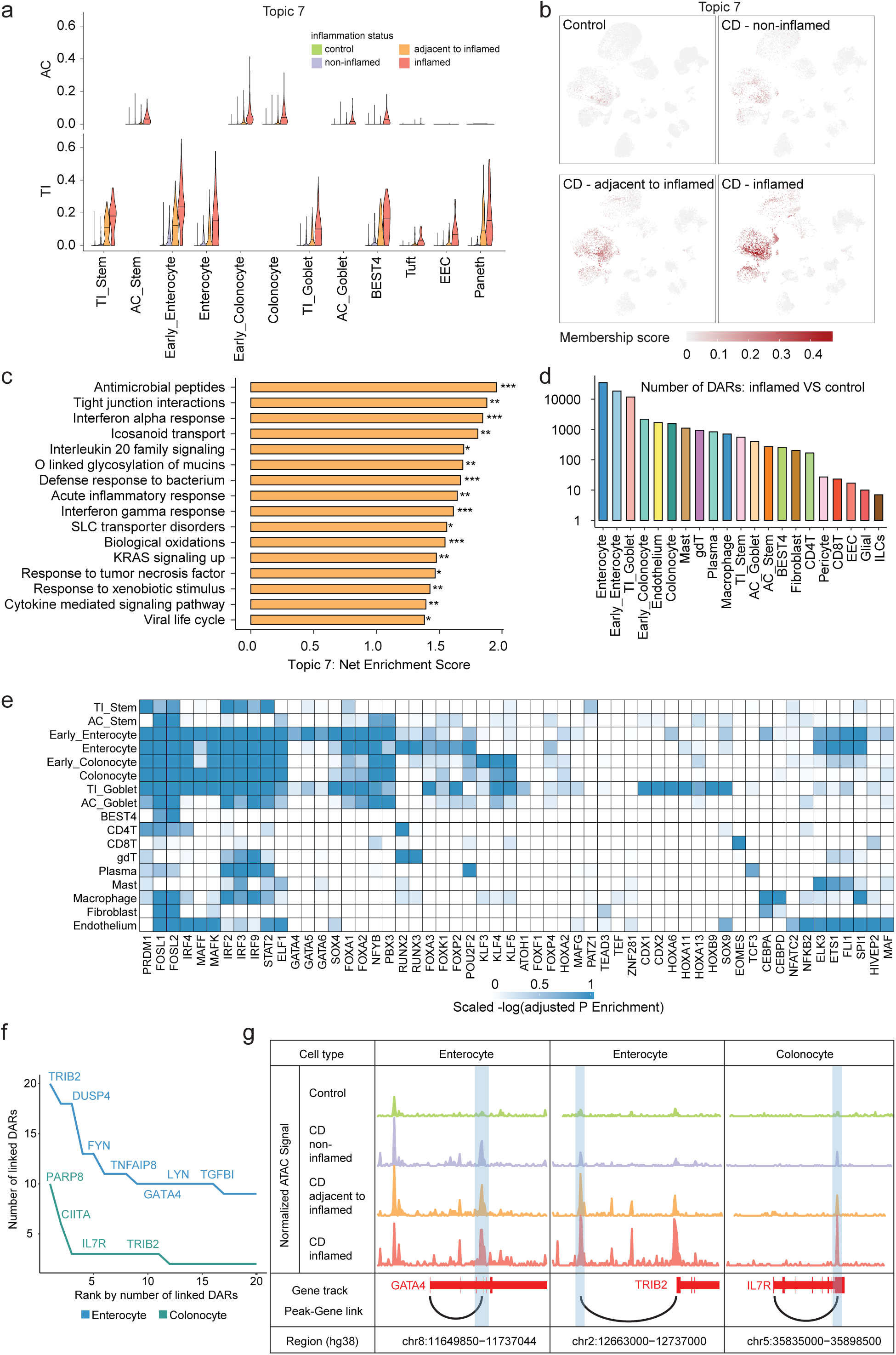
R**e**gulatory **changes associated with inflammation. a**, Proportion of topic 7 membership for each cell. Cells grouped by cell type, tissue location, and inflammation status. **b**, UMAP plots displaying proportion of topic 7 membership for each cell, split by inflammation status. **c**, Pathway enrichment analysis based on genes associated with topic 7 (***: FDR ≤ 0.001, **: FDR ≤ 0.01, *: 0.01 < FDR ≤ 0.05). **d**, Number of inflammation-associated DARs (iDARs) with increased accessibility in inflamed tissue compared with control tissue in each cell type (log fold change ≥ 0.5, FDR ≤ 0.1). **e**, Enriched TF motifs in iDARs. The -log10(adjusted p-value) for motif enrichment was capped at 5, with any values exceeding 5 being set to 5, and then scaled by the maximum value per TF. **f**, Identification of highly regulated inflammatory genes based on the number of linked iDARs. iDARs were linked to genes based on the activity-by-contact (ABC) model^71^. **g**, Examples of genomic loci containing iDARs upregulated in inflamed tissue and putative target genes obtained from ABC model.

Motif enrichment within topic 7 cCREs implicates AP-1 factors (e.g., FOSL1/2) and Interferon response factors (e.g., IRF2/3/9), as the main regulators of inflammation within epithelial cells (Fig. 3a). Putative targets are enriched for pathways associated with inflammatory processes, including interferon responses, antimicrobial peptides, and icosanoid transport (Fig. 4c). Notably, these target genes included *LCN2*, *NOS2*, *DUOX2, DMBT1, REG1A, and REG1B,* which were recently reported as characteristic of epithelial cells in patients with active CD^19^ (Supplementary Table 3). We thus identified the regulatory features associated with a broad inflammatory response in epithelial cells, suggesting that this response is orchestrated by a defined set of pro-inflammatory TFs. Topic 9 is enriched across stromal cells in inflamed samples (Fig. 3a, Extended Data Fig. 5b, c), in particular fibroblasts from AC. Topic 9 cCREs were enriched for motifs for FOSL1/2, STAT2, IRF4, but also included NFKB2.

Our topic analysis revealed inflammatory responses that are shared broadly across many cell types. To test whether inflammation was associated with cell-type-specific changes in cCRE activity we compared 21 cell types from inflamed and control samples to identify differentially accessible regions (DARs) (Methods). We focused our comparison on this contrast since we observed a strong difference between healthy control and inflamed CD samples in the topic analysis, and because endoscopic assessment of inflammation is often not sufficient to score mucosal inflammation in CD patient. Epithelial cell types yielded the most inflammation-associated DARs (iDARs, Supplementary Table 4), in particularly in the TI (enterocytes, early enterocytes, goblet cells) (Fig. 4d, methods). Epithelial iDARs are enriched in motifs for pro-inflammatory TFs (IRF, STAT2, MAFF, MAFK, FOSL1/2) and NFYB, ELF1 similar to topic 7 cCREs. This analysis also identified motifs enriched in iDARs of one or only a few cell types. For example, iDARs in early enterocytes are enriched for motifs of TFs associated with lineage specification and regeneration/injury (SOX4, FOXA1/2, GATA4/5/6). iDARs in TI Goblet cells were enriched for FOXA1/2, FOXA3 (Hickey et al) SOX4, CDX1/2, SOX9, and ATOH1. Of note, topic 17 is similarly enriched for motifs of ATOH1, SOX4 and associated with Paneth cells in TI in all conditions, this topic appears specifically enriched in TI goblet cells from inflamed samples (Fig. 3a). These observations potentially point toward plasticity in the secretory lineage in response to inflammation. In enterocytes and colonocytes we found that iDARs cluster around many genes that were previously associated with inflammatory responses and regeneration, including GATA4 (10 iDARs), TRIB2 (20 iDARs), and IL7R (3 iDARs) (Fig. 4f, g).

We identified 1702 iDARs in endothelial cells that are enriched for motifs of pro-inflammatory transcriptional regulators IRF4, STAT2, NFkB2 and AP-1 like factors, suggesting a significant inflammatory transcriptional response in this cell type which plays a key role in the regulation of cellular and molecular access to inflamed regions^44^ (Fig. 4e). The iDARs we detected in other cell types were enriched for motifs of TFs associated with inflammation or master regulators of specific cell types. For example, among immune cells, CD8+ T cells are enriched for EOMES, previously implicated in the differentiation of effector CD8+ T cells^45^, and Plasma B cells are enriched for POU2F2 (OCT2), a TF critical in Plasma cell maturation^46^. Together these data helped to infer the key regulatory TFs driving cell-type-specific regulatory programs in mucosal cells of patients with active CD.

### Testing cell-type-specific regulatory sequence motifs using chromatin accessibility QTLs

Our study generated a catalogue of cell-type-resolved cCREs which we used to infer sequence determinants and TFs regulating chromatin accessibility. To directly test the relationship between DNA sequence (e.g., TF binding motifs) and chromatin accessibility, we leveraged our cohort of 39 individuals to map caQTLs. We obtained genome sequences for each individual directly from scATAC-seq data and imputed their genotypes based on the 1000G panel (Methods). We used RASQUAL to separately map caQTLs in each of the 19 cell types that were represented by at least 20 individuals. We detected between 8533 (early colonocytes) and 21 (neutrophils) caQTLs per cell type (FDR<10%, MAF>5%) (Fig. 5a). The number of detected caQTLs depended on the total number of cells constituting the cluster, highlighting substantial differences in power between cell types (Fig. 5b). However, applying multivariate adaptive shrinkage (mash^47^) to the identified caQTLs revealed substantial sharing between related cell types (Extended Data Fig. 7h, Method), as did the correlation pattern of caQTL effect sizes between pairs of cell types (Extended Data Fig. 7i). For the subsequent analysis we used the original RASQUAL caQTLs identified for each cell type. Compared to random SNPs from our test set, RASQUAL caQTLs are enriched within promoters, they tend to be located near the center of the caPeaks (Extended Fig. 7 b-e), and they are less conserved than non-caQTL Peaks (Extended Fig. 7g). caQTL peaks have higher peak scores on average, likely reflecting higher detection power for these regions (Extended Data Fig. 7f).

**Fig. 5.**
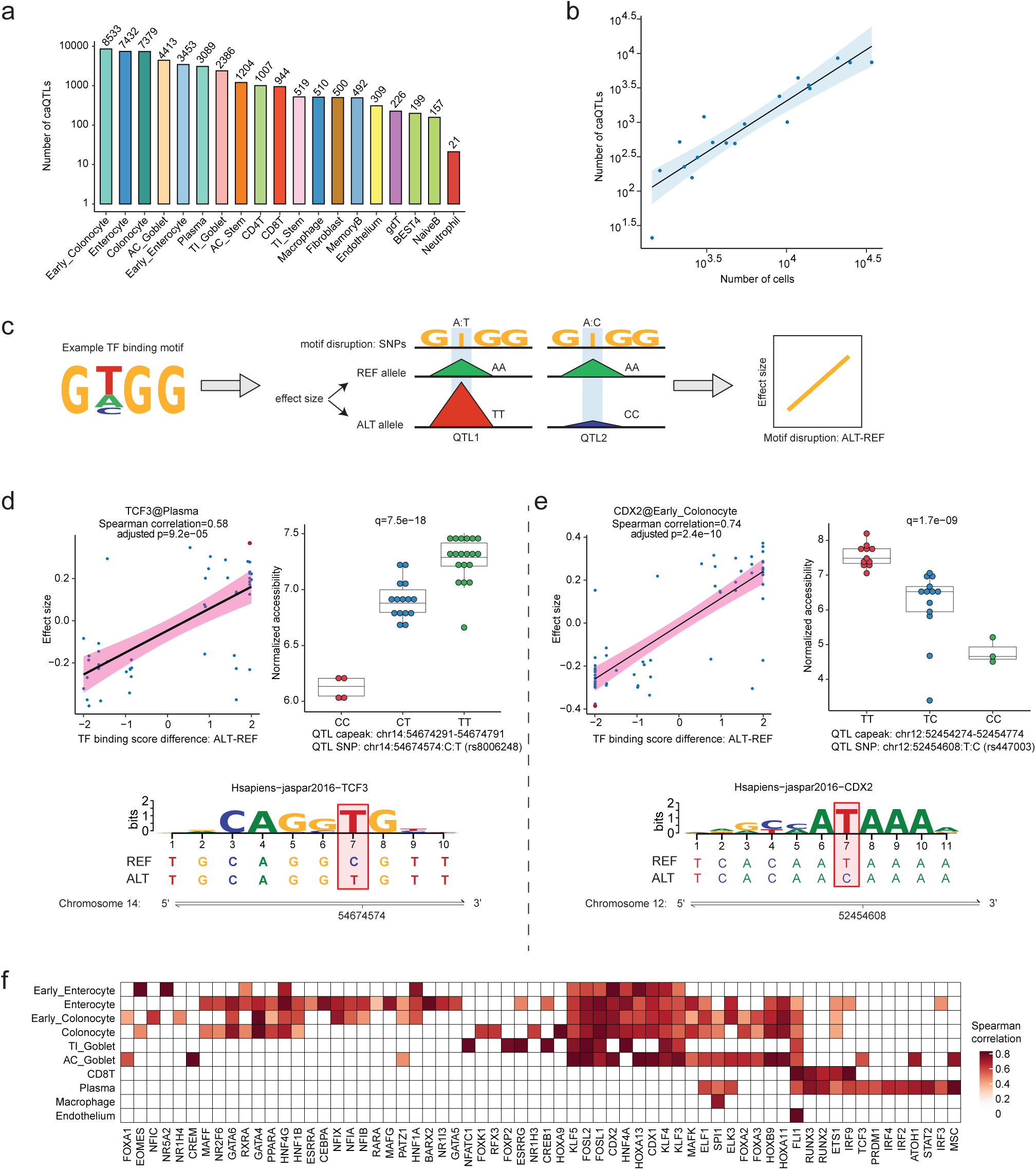
I**d**entification **of chromatin accessibility quantitative trait loci (caQTLs) in intestinal cell types. a**, Number of caQTLs identified in each cell type using RASQUAL. **b**, Relationship between the number of detected caQTLs and the number of cells in each cell type (Spearman correlation rho=0.90, p-value < 2.2e-16). **c**, Illustration of analysis to measure effects of motif disrupting SNPs on chromatin accessibility changes. In the example, differences in motif affinities for the TF at alternative alleles from QTL1 and QTL2 are associated with different accessibilities of the corresponding caPeaks. **d**, Correlation of caQTL effect sizes with motif disruption scores (MotifBreakR) for QTL SNPs predicted to disrupt TF sequence motif for TCF3. Shown are QTL SNPs located within caPeaks in plasma cells. Effect sizes and motif disruption scores were calculated relative to reference allele. Significance of association was calculated using Spearman correlation. Box plots show peak accessibilities in different genotypes illustrating the effect of a specific SNP (highlighted in red in the regression plot). Genome tracks in the bottom show reference and alternative alleles at the QTL SNP site. **e**, Same as in **d**, showing chromatin accessibility at CDX2 motifs in early colonocyte clusters. **f**, Heatmap of Spearman correlation coefficients between QTL motif breaking scores and effect sizes for all TF-cell type pairs. Coefficients are only displayed for significant correlations (FDR <0.05).

In principle, caQTLs allow us to detect TFs that control chromatin accessibility by observing the relationship between TF motif disruption and changes in chromatin accessibility^48,49^. For each cell type, we obtained TF motif instances that overlapped with a caQTL and were predicted to be significantly altered by that SNP (Methods). We then tested the correlation between caQTL effect size and motif disruption score separately for each set of motifs and retained motifs with significant correlation in at least one cell type (Fig. 5c). We observed a significant correlation between motif disruption and chromatin accessibility in at least one cell type for 64 TF motifs (adjusted p-value ≤ 0.05, Spearman’s correlation coefficient) (Fig. 5d-f). For these specific sites, we found strong evidence that disruption of binding motifs of key lineage-specific TFs is associated with reduction in chromatin accessibility at these cCREs; for example, TCF3 in Plasma cells (Fig. 5d) and CDX2 in colonocytes (Fig. 5e). While the caQTL analysis was performed across all samples regardless of disease status, we detected a significant correlation for IRF9 and IRF3 in enterocytes, providing additional evidence that these proinflammatory TFs regulate chromatin accessibility at cCREs in this cell type (Fig. 5f). These results were robust to the selection of SNP sets (Extended Data Fig 7j). Our analyses thus demonstrate that integration of inter-individual genetic variation into cellular atlases can be used to causally link cell-type-specific sequence determinants to chromatin accessibility and to validate the functional impact of motifs identified through cell-type-specific motif enrichment.

### Cell-type-specific enrichment of IBD risk

The CD gut cell atlas provided us with the unique opportunity to ask which cell types and which context-specific regulatory programs might mediate IBD risk. We used stratified LD-score regression (sLDSC^50^) to assess heritability enrichment from GWAS summary statistics for IBD and its subtypes CD and UC in cell-type- and topic-specific chromatin accessibility^14^. As reference traits we also included Celiac disease^51^, Rheumatoid arthritis (RA)^52^, Alzheimer’s disease^53^, Parkinson’s disease^54,55^, Type 1 Diabetes (T1D)^56^, Type 2 Diabetes (T2D)^57^, and Body Mass Index (BMI)^58,59^.

We found ten of the 29 cell types to be associated with enriched IBD risk based on sLDSC (ρ* > 0, FDR < 10%), all of which represent immune cells (Fig. 6a). T cells (CD4, CD8, ψ8T) and, to a lesser degree, NK cells are enriched for IBD, CD, and UC risk, as well as for Celiacs disease, RA, and T1D. GC, Naïve B, and memory B cells are less strongly enriched for IBD/CD/UC and show higher enrichment for RA and T1D, likely reflecting the humoral component of these autoimmune diseases^60,61^. Of note, we found that risk for CD, but not risk for UC, is strongly enriched in innate immune cells (macrophages, DC, neutrophils). This suggests that known differences in disease etiology and genetic architecture between UC and CD are partly caused by the cell-type-specific contribution to disease risk^62,63^.

**Fig. 6.**
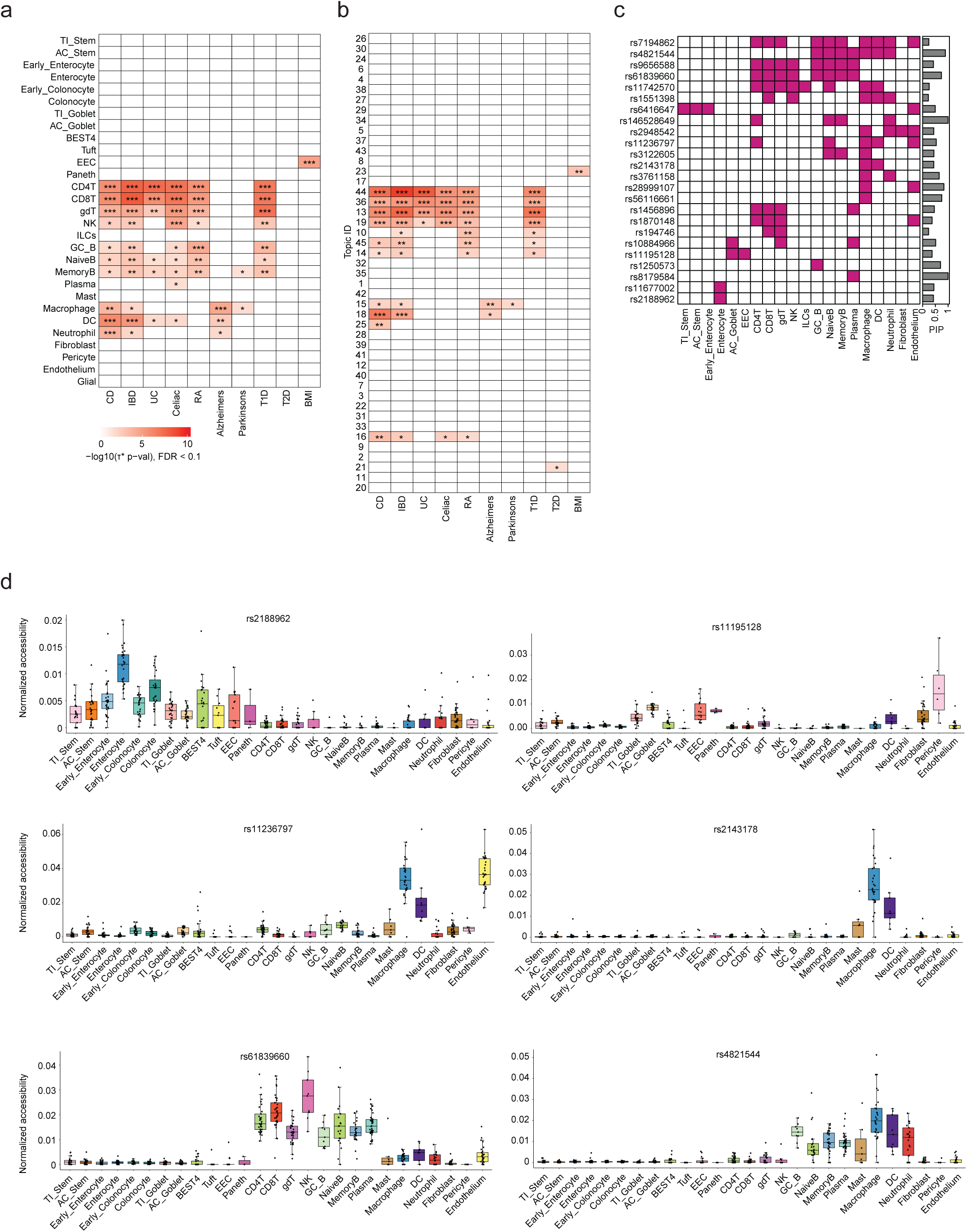
C**e**ll**-type-specific genetic risk for IBD. a**, Enrichment of trait heritability in cell-type- specific peaks using s-LDSC. Shown are cell-type-specific -log10 p-values for positive τ^∗^ evaluating autoimmune and control traits. Only statistically significant results (FDR<10%) are colored. IBD=Inflammatory Bowel Disease, UC=Ulcerative Disease, RA=Rheumatoid Arthritis, T1D=Type 1 Diabetes, T2D=Type 2 Diabetes, BMI=Body Mass Index. **b**, Topic-specific -log10 p- values for positive !^∗^ evaluating autoimmune and control traits. Only statistically significant results (FDR<10%) are colored. **c**, Shown are 24 SNPs were prioritized based on posterior inclusion probabilities (PIPs ≥ 0.2) and overlap with cell-type-specific cCREs. Ruby red color indicates that the fine-mapped SNP is located within cell-type-specific cCREs. PIPs for each SNP are shown as bar graphs on the right of the heatmaps. **d**, Distribution of normalized chromatin accessibility of pseudo-bulk cell groups per cell type per individual in the cCRE overlapping selected finemapped SNP.

**Fig. 7.**
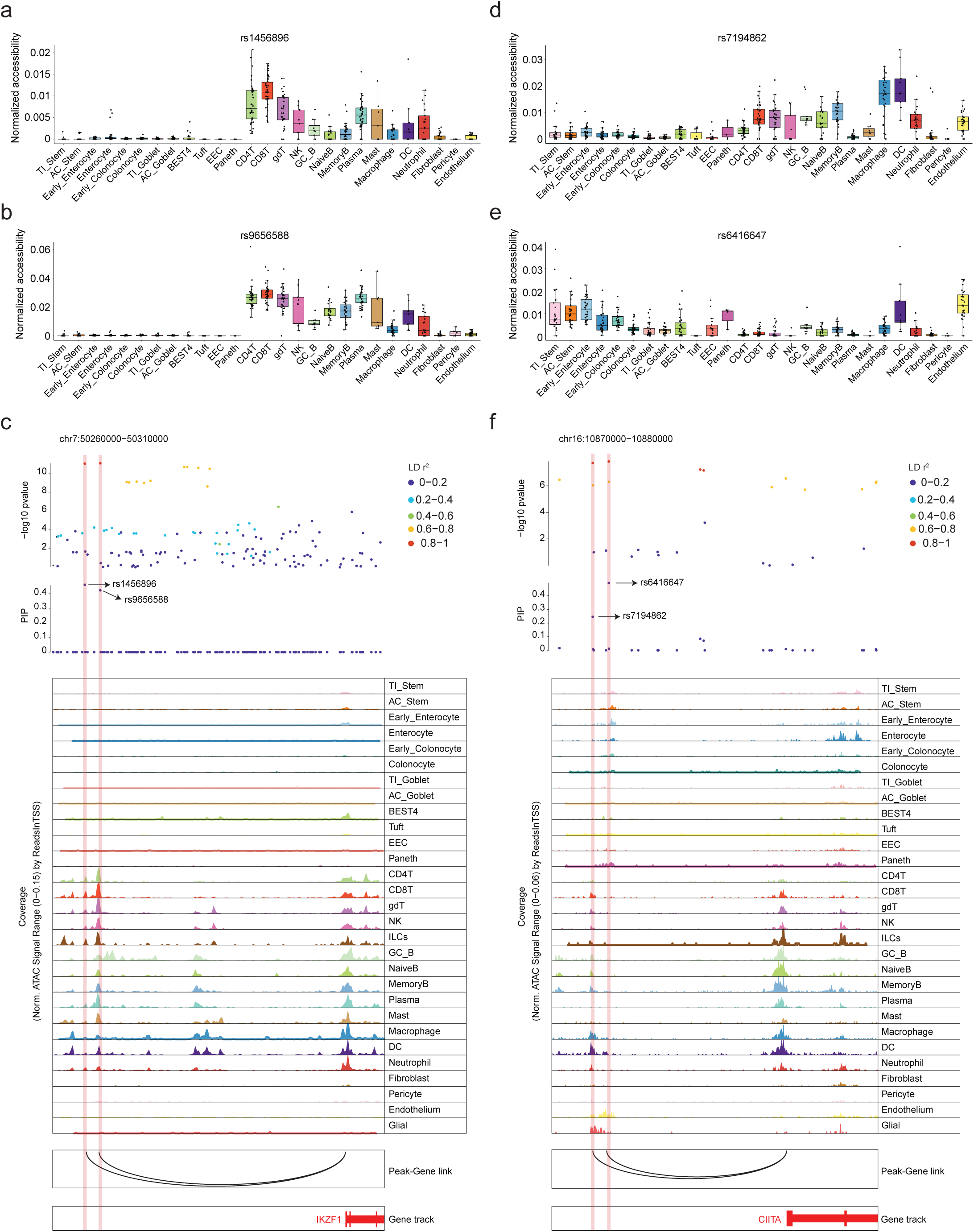
C**a**ndidate **functional risk variant prioritization. a**, **b**, **d**, **e**, Distribution of normalized chromatin accessibility of pseudo-bulk cell groups per cell type per individual in the cCRE overlapping the finemapped SNP. **c**, Fine-mapped CD risk SNPs rs1456896 and rs9656588 are from the same fine-mapped credible set. The top track represents the -log10 p-value of SNPs from CD GWAS, with color representing LD with the lead SNP. The middle track shows PIPs from fine-mapping results. In the bottom track, IKZF1 was linked to the cCREs by the co-activity of gene expression and chromatin accessibility. **f**, Fine-mapped IBD risk SNPs rs6416647 and rs7194862 are from the same fine-mapped credible set. In the bottom track, CIITA was linked to the cCREs through the ABC model.

In contrast, none of the epithelial or stromal cell populations are enriched for IBD risk. However, we detected strong enrichment for BMI heritability in EEC-specific peak regions, confirming a previous observation^31^ and suggesting that part of the genetic contribution to BMI is mediated via EECs. We next used topic-associated peaks to assess association of contexts with IBD risk. Our results confirmed the association of specific immune cell types with IBD risk and the difference in CD and UC risk heritability in innate immune cells (topics 15, 18, 25). Similarly, we found no evidence for enrichment of IBD risk in epithelial (topics 3-7, 17, 22-24, 26, 27, 29-31, 33, 34, 37, 38, 40) or stromal cells (topics 9, 28, 39) using topic cCREs (Fig. 6b).

In contrast to cell-type-cluster-based enrichment, topic analysis also captured inflammation-associated cCREs. However, topics 7 and 9, corresponding to epithelial and stromal cells, respectively, were not enriched for IBD, CD or UC heritability. Together these analyses point to immune cell types with large contributions, while suggesting that loci mediating strong inflammatory responses in the epithelium and stromal cells, have a more limited contribution to overall genetic IBD risk. Our analysis provides an example of how context-specific epigenetic data and genetic data can be combined to disentangle (cell-type-specific) genetic effects from their cellular consequences in complex diseases.

### Cell-type-resolved cCREs enable prioritization of candidate regulatory CD variants across mucosal cell types

In contrast to the results of our heritability enrichment analysis, prior data implicated genes with epithelial function in IBD and its subtypes, including *CIITA* and *SLC22A4*^64–66^. sLDSC does not test for cell type specificity of individual loci, but instead obtains a genome-wide estimate of the focal heritability enrichment for a given annotation across all loci (ρ*)^50,67,68^. Thus, sLDSC likely misses cell types for which the contribution to overall genetic risk comes from a small number of loci. To link risk loci to cell-type-specific activity, we fine-mapped the CD GWAS signal and obtained posterior inclusion probabilities (PIPs) for variants in 91 loci (Methods). We noted that variants with high PIP are enriched within cCREs (enrichment=3.5-fold, p-value=0.0096, Fisher’s exact test; Methods, Extended Data Fig 8a, b), and in 24% of loci (20/85) at least 50% of the total PIP is associated with cCREs (Extended Data Fig 8c). We identified 29 variants in 24 loci (25 credible sets) with a PIP ≥ 0.2 that fall within cCREs (Table 1). Where possible, we nominated a putative target gene for these variants using co-accessibility^34,69^, published Activity-by-contact (ABC) scores^70,71^, or distance as criteria. Out of the 29 variants, 24 overlap with at least one cell-type-specific cCRE (Fig. 6c). As expected, most of these regions are accessible in adaptive and innate immune cells. However, we identify 10 variants overlapping a region identified as specific to at least one epithelial or stromal cell cluster, implicating these cell types in mediating IBD risk (Fig. 6c). For example, rs2188962 (PIP=0.42) is located within a region broadly accessible in epithelial cells, most strongly in mature enterocytes and colonocytes (Fig. 6d), and rs11195128 falls into regions accessible in secretory epithelial cells (Fig. 6d). Additional prioritized SNPs are located in cCREs accessible in T and B cells, Macrophages and DCs, as well as endothelial cells. Several of the prioritized SNPs have been implicated in IBD and validated through functional studies (e.g., rs11236797 and rs61839660^72^, Table 1). Together, these analyses implicated a range of immune and non-immune cells in the intestinal mucosa in mediating CD risk through regulatory variants.

**Table 1.** Prioritized risk SNPs. Risk SNPs with a PIP greater than 0.2 that overlapped with a cCRE were listed. The target gene was first linked to the SNP based on co-activity. If no genes showed co-activity with the overlapping peak, the ABC-Max method was used to assign the associated gene. In cases where neither approach was applicable, the nearest gene was assigned. The lineage specificity is summarized from the cell type specificity of the overlapped cCRE. The iDAR column specifies whether the overlapped cCRE is classified as an iDAR.

| rsID | PIP | cCRE | Associated gene | Link method | Lineage specificity | iDAR | Known IBD risk Gene [PMID] |
| --- | --- | --- | --- | --- | --- | --- | --- |
| rs8179584 | 1 | chr2:172558327-172558827 | PDK1 | ABC | Immune |  | [38103512] |
| rs146528649 | 1 | chr16:50626815-50627315 | SNX20 | Co-activity | Immune |  | [32410873] |
| rs2284553 | 0.97 | chr21:33404169-33404669 | IFNGR2 | ABC |  |  | [31566580] |
| rs4821544 | 0.89 | chr22:36862189-36862689 | NCF4, CSF2RB | Co-activity | Immune |  | [18580884, 27377463] |
| rs28999107 | 0.83 | chr12:6383870-6384370 | PLEKHG6, TNFRSF1A, LTBR | Co-activity | Immune, Stromal |  | [30815272, 33568785] |
| rs56116661 | 0.75 | chr3:188683082-188683582 | LPP-AS1 | ABC | Immune |  |  |
| rs61839660 | 0.74 | chr10:6052545-6053045 | IL2RA | ABC | Immune | ✓ | [34226674] |
| rs1250573 | 0.7 | chr10:79282308-79282808 | ZMIZ1 | Nearest | Immune |  | [30402962] |
| rs3761158 | 0.65 | chr20:46006257-46006757 | MMP9 | ABC | Immune | ✓ | [27542125] |
| rs11236797 | 0.53 | chr11:76588423-76588923 | LRRC32 | Co-activity | Immune, Stromal | ✓ | [32499651] |
| rs10884966 | 0.51 | chr10:110425817-110426317 | DUSP5 | ABC | Epithelial, Immune |  | [26479789] |
| rs6416647 | 0.49 | chr16:10871644-10872144 | CIITA | ABC | Epithelial, Stromal | ✓ | [35275975] |
| rs1456896 | 0.46 | chr7:50264832-50265332 | IKZF1 | Co-activity | Immune |  | [31057532] |
| rs3122605 | 0.44 | chr1:206781236-206781736 | IL10 | Nearest | Immune |  | [28496525] |
| rs11677002 | 0.44 | chr2:28391530-28392030 | FLJ31356 | ABC | Epithelial |  |  |
| rs2143178 | 0.44 | chr22:39264648-39265148 | PDGFB | ABC | Immune |  | [37995775] |
| rs2948542 | 0.44 | chr17:27529222-27529722 | KSR1 | ABC | Immune, Stromal |  | [34758847] |
| rs2188962 | 0.42 | chr5:132434879-132435379 | P4HA2, SLC22A4, SLC22A5 | Co-activity | Epithelial |  | [38944815, 15503241] |
| rs9656588 | 0.42 | chr7:50266738-50267238 | IKZF1 | Co-activity | Immune |  | [31057532] |
| rs11195128 | 0.37 | chr10:110426330-110426830 | DUSP5 | ABC | Epithelial |  | [26479789] |
| rs1870148 | 0.37 | chr10:80511347-80511847 | LOC102723703 | ABC | Immune, Stromal |  |  |
| rs4807569 | 0.32 | chr19:1122945-1123445 | MIDN, EFNA2 | Co-activity |  |  | [37782162] |
| rs2031754 | 0.29 | chr9:4965364-4965864 | JAK2 | Nearest |  | ✓ | [32203403] |
| rs11742570 | 0.26 | chr5:40410171-40410671 | PTGER4 | ABC | Immune |  | [36155972] |
| rs7194862 | 0.25 | chr16:10871018-10871518 | CIITA | ABC | Immune, Stromal | ✓ | [35275975] |
| rs194746 | 0.23 | chr14:68815969-68816469 | ZFP36L1 | ABC | Immune |  | [30402962] |
| rs7608697 | 0.23 | chr2:60977210-60977710 | PUS10 | Nearest |  |  | [21298027] |
| rs2024092 | 0.23 | chr19:1123791-1124291 | SBNO2 | Nearest |  | ✓ | [38806456] |
| rs1551398 | 0.22 | chr8:125527564-125528064 | TRIB1 | Nearest | Immune | ✓ | [28187197] |

Of note, we also identified two loci in which two prioritized non-coding variants from the same credible set overlapped separate regulatory elements. In the first locus, rs1456896 (PIP = 0.46) and rs9656588 (PIP = 0.42) which are separated by 2.3kb, overlap adjacent cCREs, both of which are linked to IKZF1 by co-activity (Fig. 7a, b, c). Both cCREs display similar accessibility patterns across cell type clusters, with the highest accessibility in T cells. In the second locus, rs7194862 (PIP =0.25) and rs6416647 (PIP =0.49) are separated by 482 bp and located in cCREs linked to *CIITA*^73^ (Fig. 7d, e, f), a transcriptional co-activator of *MHCI* and *MHCII* genes^73^. In this case, chromatin accessibility of the two cCREs harboring these variants differs significantly across cell types. While rs7194862 is located in a cCRE accessible in macrophages, DC, T cells and B cells and endothelial cells, rs6416647 is accessible in stem cells and absorptive epithelial cells (and Paneth cells), DCs and endothelial cells (Fig. 7d, e, f). These two examples highlight the potential contribution of multiple non-coding SNPs to CD risk within a single locus and mediation of this risk by different cell types.

## Discussion

We assembled an atlas of cell-type-resolved chromatin accessibility within the intestinal mucosa of adult CD patients and healthy controls and used it to characterize cell-type- and context-specific gene regulatory programs across disease-relevant cell types and states. Our work contextualizes findings from previous scRNA-seq studies, which identified cell-type-specific and inflammation-associated gene expression signatures in CD^18,19^. In particular, we provided direct insights into the cCREs and TFs that define the regulatory landscape of the intestinal mucosa.

Cell-type-specific TF motif enrichment is a common approach for nominating putative regulatory elements^34,74^. If gene expression data is available, the enriched motifs can then be integrated with transcript levels for the cognate TFs. However, such approaches do not assess whether the enriched motifs directly contribute to establishing or maintaining chromatin accessibility in vivo. Our study, which included 71 tissue samples from 39 individuals provided us with the unique opportunity to functionally test the effects of TF motifs in putative cCREs across a range of disease-relevant cell types and contexts. By examining the overlap between caQTLs and putative regulatory SNPs in cCREs, we identified 64 TF motifs that directly affect chromatin accessibility in the intestinal mucosa - even with our moderate sample size. As cohort sizes continue to increase, we expect that similar approaches will become a standard step in analyzing data from epigenomic atlases and to predict regulatory effects of GWAS SNPs^29^.

scATAC-seq has been used across a wide range of tissue contexts to identify cell types mediating genetic risk for a variety of traits^68^. Similar to previous studies in intestinal tissues, we observed strong enrichment for IBD and CD risk in both in adaptive and innate immune cells^31,33^. while UC shares genetic architecture with CD, we found no or little enrichment of UC risk in innate immune cells, reflecting known differences between UC and CD and demonstrating both the sensitivity and specificity of our approach^63^. Unexpectedly, we found that neither epithelial nor stromal cells are enriched for CD risk, nor are inflammation-associated cCREs. This observation is at odds with the evidence for GWAS loci implicating genes with epithelial function^9,17,33,75^. As sLDSC measures enrichment scores across multiple loci (i.e., including but not limited to cCREs), it likely produced conservative estimates of heritability enrichment in cCREs. The combination of context-specific epigenomic data which detects strong inflammation associated changes in epithelial cell types and cell-type-specific risk primarily associated with immune cells prompts us to speculate that we are separating primary genetic risk from secondary effects in CD.

Fine-mapped variants within cCREs have previously been used to identify putative causal non-coding variants with direct effects on gene regulation^21,29^. Among the 29 putative regulatory variants of CD risk, we found 24 falling into cCREs with cell-type-specific accessibility patterns. Of note, several of these variants are located in cCREs with epithelial specific chromatin accessibility. Intriguingly, we also observed cases in which multiple variants in LD with similar PIP scores fall into adjacent regulatory elements, including one example where adjacent elements implicate distinct cell types. Together, these prioritized variants highlight the gene regulatory complexity underlying CD and provide evidence for a broad involvement of both immune and non- immune cell types mediating CD risk.

## Data availability

All data produced in the present study will be shared upon publication.

## Acknowledgement

This work was supported through grants from the National Institutes of Health R35GM142986 (to S.P.). This publication is part of the Gut Cell Atlas Crohn’s Disease Consortium funded by The Leona M. and Harry B. Helmsley Charitable Trust and is supported by a grant from Helmsley to The University of Chicago (https://helmsleytrust.org/gut-cell-atlas/). This work was completed in part with resources provided by the University of Chicago Research Computing Center. We thank Matthew Stephens for helpful discussion of the analysis, Katie Aracena for the discussion of GWAS data preprocessing and SNP rsID annotation, and Natalia Gonzales for assistance with editing.

## Author contributions

Y.Z. performed data analysis. R.Z. generated data. Z.M. and P.C. contributed to data analysis. X.Z., B.X., K.L., X.H., A.W.D., X.L. and E.B.C. contributed to data interpretation. C.M.C. and J.K. contributed to sample collection, processing and experiments. Y. Z. and S.P. wrote the manuscript and created figures. S.P. conceptualized the study. A.B. and S.P. supervised the study.

## Methods

### Ethics statement

This study protocol (15573A) was approved by the Institutional Review Board of The University of Chicago. Gastroenterologists collected samples and clinical data from patients diagnosed with Crohn’s Disease (CD) and non-IBD control. Mucosal samples were collected during routine endoscopic screening in healthy controls, or during diagnosis or follow-up colonoscopy in CD patients. All patients provided signed consent for research use of their mucosal biopsies and storage of their demographic data. Samples, clinical metadata and demographic data were collected by clinical research coordinators and provided anonymously to the research team. This study complied with all relevant ethical regulations.

### Patient information

Non-IBD controls were recruited by the time of routine colonoscopy. Non-IBD controls were free of a history of IBD, an auto-immune disease, an ongoing infection in intestines, a history of Familial adenomatous polyposis, a history of primary digestive tract malignancies, or a history of metastatic malignancies in digestive tract. CD patients were recruited at the diagnosis or follow-up colonoscopy. Those patients with prior clinical diagnosis of CD, were reported either in clinical remission or with active disease. Patient and sample metadata are provided in Supplementary Table 1.

### Tissue biopsy sample collection

We aimed to obtain samples from two anatomic locations, terminal ileum (TI) and ascending colon (AC), and in the case of CD patients with active inflammation from inflamed sites and adjacent regions without macroscopic inflammation. The specific sampling regions within anatomic locations were determined by the gastroenterologists performing the colonoscopy. The number of pinches were limited to 6 per individual site and 12 per patient. CD patients provided up to twelve biopsies, including up to two regions (TI and AC), and three different endoscopic diagnoses (inflamed, adjacent to inflamed, noninflamed)^76^. In most cases with active inflammation, CD patients only provided biopsies from a single anatomic location (i.e. AC or TI). Non-IBD controls provided up to six biopsies from each region (TI and AC). Mucosal linings were obtained using endoscopic biopsy forceps following standardized care. Mucosal tissues after biopsy were immersed in iced cold Advanced DMEM/F-12 [Thermo Fisher 12634010] in a 1.7 mL eppendorf tube and transferred in a cooler. The maximum wait time between biopsy and sample processing was two hours.

### Sample dissociation and nuclei extraction

Single cells from mucosa were obtained following a modified protocol of previous studies^38,77,78^. Mucosas were rinsed and agitated in 1.5 ml ice-cold PBS [Thermo Fisher 10010023] until the supernatant looks clear. Mucosa from the same regions and same endoscopic diagnosis were minced by iris scissors. Minced tissues were immersed in 2 ml warm (37°C) detachment solution (HBSS Ca/Mg-Free [Thermo Fisher 14175095], 5 mM EGTA [Fisher Scientific 50-255-956], 10 mM HEPES [Thermo Fisher 15630080], and 10% FBS [Gemini Bio 100-106]). The tissues were incubated at 37°C for 15 minutes with end-over-end rotation, followed by 10-minute cool-down on ice. The tissues were agitated (shake 20 times) to separate epithelium from lamina propria. Supernatant containing epithelial sheets, was collected and spun down at 300g for 5 minutes at 4°C. The remnant tissue was rinsed in ice-cold PBS twice. The epithelium was incubated in 2 ml TrypLE express [Thermo Fisher 12605010], at 37°C for 5 minutes with rotation. The remaining tissues were incubated in 2 ml enzyme solution (HBSS Ca/Mg-Free [Thermo Fisher 14175095], 10 mM HEPES [Thermo Fisher 15630080], 200 μg/ml Liberase TM [Millipore Sigma 5401127001] and DNase I [Gold Bio D-301-100]) at 37°C for 20 minutes with rotation. After enzymatic digestion, epithelium and lamina propria were triturated with a P1000 pipette until no tissue chunks is visible. Cells were filtered through a 40 μM cell strainer [Falcon/VWR 21008–949]. The filtrated cells from both digestions were pooled and spun down at 300 g for 5 minutes at 4°C. Cell pellets were resuspended in 200 μl cold wash buffer (HBSS Ca/Mg-Free [Thermo Fisher 14175095] and 10% FBS [Gemini Bio 100-106]). Cell suspension was then diluted by 2 ml RBC lysis buffer [Miltenyi Biotec 130-094-183] and incubated for 3 minutes at room temperature. Cells were then spun at 300g for 5 minutes at 4°C. Cell pellets were resuspended in 200 μl suspension buffer (PBS and 0.04% BSA [Miltenyi Biotec 130-091-376]) and assessed viability with Trypan Blue [Thermo Fisher 15250061] and counted. Only cell suspensions with > 90% viability were used for downstream single cell assays. The detailed processing is fully described here^79^. After cells were partitioned in the scRNA-seq assay, the remaining cells were pelleted again by spinning at 500g for 5 minutes at 4°C. After the removal of supernatant, 100 μl lysis buffer (CG000169 Rev E, 10x Genomics) was added to each pellet, followed by gently pipetting. The suspension was incubated on ice for 4 minutes and diluted by 1 ml wash buffer (CG000169 Rev E). Nuclei were spun down at 500g for 5 minutes at 4°C. After resuspending in 1 ml wash buffer, nuclei were pelleted again. The nuclei were suspended in 20 μl 1x Nuclei buffer (10x Genomics, PN-2000153/2000207) and assessed for density with Trypan Blue. The final density of nuclei was adjusted to 1.6 million per mL with 1x Nuclei buffer.

### scATAC library preparation and sequencing

Single nuclei were isolated from single cell suspensions following the manufacturer’s protocol for the Chromium Single cell ATAC (v1.1). Briefly, single nuclei were transposed before loading into chip H. Transposed nuclei were partitioned into GEMS in Chromium controller. GEMs were incubated for barcoding, followed by indexed amplification. Approximately 8,000 nuclei were loaded per channel to target recovery of 5,000 nuclei. Libraries were sequenced on Illumina Nextseq or Novaseq platforms at the Genomics Core Facility at The University of Chicago.

### scRNA-seq data preprocessing and analysis

scRNA-seq raw data were obtained from Zhou et al. and processed using Cell Ranger (v7.0.1) with the 10x human transcriptome GRCh38-2020-A as the reference. SoupX was used with default parameters to estimate and remove cell free mRNA contamination for each sample individually^80^. We used the graph based clustering results from Cell Ranger output to estimate the contamination fraction. Downstream analysis was performed in R (v4.1.0) and Seurat^81–85^ (v4.2.0). Individual cells matching the following criteria were retained for analysis: more than 500 UMIs and fewer than 20,000 UMIs; more than 200 unique genes and fewer than 6,000 unique genes; less than 50% of reads mapping to mitochondrial genes; less than 40% of reads mapping to ribosomal genes; and a ‘log10GenesPerUMI’ score higher than 0.7 (to exclude low-complexity cell types such as red blood cells). Doublet scores were calculated using Scrublet^86^ (v0.2.3), and cells with score of 0.2 or above were considered doublets and removed. The cells exhibiting co-expression of different lineage makers (epithelial: EPCAM, T cell: CD3D, B cell: MS4A1, CD79A, IGHA1, myeloid: TYROBP, CD14, CPA3, stromal: IGFBP7, COL3A1, endothelial: PECAM1) were also excluded as doublets. Genes expressed in fewer than 3 cells per sample were filtered out. Transcript counts for each cell were normalized and log-transformed. Cell cycle scores (the difference between the G2M and S phase scores) were calculated with cell cycle genes provided in Seurat (v4.2.0) using the CellCycleScoring Function. Cell cycle scores, the percentage of mitochondrial reads, and the number of unique molecular identifiers (UMIs) were regressed out before data scaling.

We identified the 2,000 most highly variable genes (HVG) by modeling the mean-variance relationship for each gene across all cells and used the count matrix restricted to these HVGs as input for principal component analysis (PCA). We selected the top 30 principal components for subsequent clustering and Uniform Manifold Approximation and Projection (UMAP) embedding^87,88^. Cell clustering was performed using the Louvain algorithm (resolution 0.3–1.5) for modularity optimization with the kNN graph as input^89^.

The union dataset was divided into three different lineages (epithelial, immune and stromal) based on clustering analysis and marker gene expression. Cells from each lineage were subset for further analysis and we repeated the process of dimensionality reduction, Louvain clustering and UMAP visualization for each lineage as described above. We filtered out clusters without specific markers, doublet clusters, and cells likely not belonging to the specific lineage through iterative clustering and annotation procedures. Cell identities for subtypes within each lineage were assigned based on previously reported marker genes^36^, with a summary provided in Extended Data Fig. 2.

### scATAC data preprocessing

scATAC-seq raw sequencing data were processed using the Cell Ranger ATAC v2.0.0, and the 10x human genome GRCh38-2020-A-2.0.0 as the reference. We used cellranger-atac count to generate scATAC fragment files from fastq files obtained from sequencing core facility. Further filtering of raw data was conducted within the ArchR (v1.0.3) framework^34^. We kept high quality cells for downstream analysis (minimum fragments per cell = 5,000, minimum TSS enrichment score = 6). Doublet scores were calculated using addDoubletScores function (nTrials = 10, dimsToUse = 2:50), and cells predicted to be doublets were removed in downstream analysis (a maximum of 10% of the cells are predicted to be doublets).

### scATAC data analysis

We followed the standard ArchR workflow to process the fragment files into a 500 bp tile matrix of fragment counts and generated a gene score matrix using the default gene score model (model 42). Latent semantic indexing (LSI) was performed on the tile matrix, retaining the top 100 LSI vectors by applying the ‘addIterativeLSI’ function with six iterations and five different clustering resolutions (0.5, 1, 1.5, 2, and 2), and up to 150,000 variable features. The first LSI vector was excluded due to its high correlation with sequencing depth. Cell clusters were identified using the ‘addClusters’ function, and clusters displaying high gene scores for multiple markers representing different lineages (epithelial, immune, stromal) were labeled as doublets and removed, similar with scRNA-seq annotation. We then divided the cell type clusters into the three lineages and performed iterative dimensionality reduction, clustering, and filtering within each lineage. We filtered out the clusters without a specific marker, doublet clusters and cells identified as not belonging to the correct lineage during each iteration.

Within each lineage, the scRNA-seq gene expression matrix was integrated with the scATAC-seq gene score matrix using ArchR’s ‘addGeneIntegrationMatrix’ function. Corresponding cells across datasets were matched using Seurat’s mutual nearest neighbors algorithm, facilitating the transfer of labels from scRNA-seq to scATAC-seq data. The scATAC-seq labels were further refined based on the distribution of marker gene scores.

### Accessible chromatin peak calling

To obtain regions of accessible chromatin (‘peaks”) in each cell type cluster, we followed the peak calling workflow introduced in ArchR. Cells within each cell type were merged to generate pseudo- bulk replicates using the addGroupCoverages function. The addReproduciblePeakSet function was then utilized to call peaks for each pseudo-bulk replicate using MACS2. Within each cell type, peak sets from all pseudo-bulk replicates were combined using the iterative overlap peak merging procedure, resulting in a merged peak set for that cell type. Finally, peak sets from all cell types were merged to create a union peak set using the same procedure. The peaks were classified into four categories based on the summit positions: promoters or promoter proximal regions (2000 bp upstream and 100 bp downstream of a TSS), intronic, exonic, distal (>2,000 bp from TSS and intergenic).

### Cell-type-specific differential accessible region (DAR) analysis

To identify cell-type specific marker peaks, a single-cell insertion count matrix was created using the function addPeakMatrix in ArchR. Differentially accessible regions (DARs) for each cell type, relative to the others, were identified using Wilcoxon rank-sum tests among cells from all healthy individuals (FDR ≤ 0.1, logFC ≥ 0.5), excluding the effects of inflammation. To control for technical variation, the single cell TSS enrichment score and number of fragments per cell were considered when selecting a matched null group. The DARs identified in each cell type were defined as cell type specific peak set and provided in Supplementary Table 4.

### Pairwise DAR identification between different inflammation conditions

We conducted pseudo-bulk based differential chromatin accessibility tests following dreamlet (v0.99.6) workflow^90^. Raw counts were first aggregated across cells within each sample and cell type cluster. The pseudobulk counts within each cell cluster were normalized using the voomWithDreamWeights function to account for patient-specific random effects. A regression model was subsequently fitted for each peak and cell cluster to test differential accessibility between pairs of conditions (control, noninflamed, adjacent to inflamed, inflamed), with sex and age included as covariates. iDARs were selected based on a log-fold change greater than 0.5 and an adjusted p-value less than 0.1, when comparing inflamed samples with control samples. The cell type specific iDARs were provided in Supplementary Table 4.

### Topic modeling and pathway enrichment

We used FastTopics^42,43^ to fit topic models to the scATAC peak matrix. To reduce computational burden, we subsampled the binarized union scATAC peak matrix by including a maximum of 100 cells per cell type per sample. This also balanced cell counts from different lineages and helped to avoid obtaining a large number of topics associated with epithelial cells at the expense of interpretable topics from other lineages. We fitted Possion non-negative matrix factorizations (NMF) and topic models for the subsampled peak matrix using the fit_poisson_nmf and poisson2multinom functions available in fastTopics^42,43^ (v0.6-159). The factors and loadings were initialized uniformly at random and then updated through the expectation-maximization (EM) algorithm for the first 80 iterations. Subsequently, updates were made using the sequential coordinate descent (SCD) algorithm until convergence, achieved after 670 iterations.

To select the number of topics (k), we tested topic resolutions k=10 to k=50 in increments of 5 aiming to balance the desire for high resolution with the ability to interpret the results. We found only a few additional interpretable topics added between 40 to 50 and chose k=45 for subsequent analysis. In many cases additional topics were strongly correlated with previously discovered ones, providing similar sets of peaks, enriched motifs, and putative target genes. To ensure that we did not systematically miss regulatory topics in this combined analysis, we repeated this process for each individual lineage (epithelium, immune, stromal). We increased the number of cells per sample and cell type cluster to 300. Interpretable topics uncovered in individual lineages mapped well to topics uncovered using the union set of all cells. We also assessed whether any of the additional topics associated with new features (e.g. location or inflammation) obtained at higher resolution were enriched for CD heritability but found no new associations. .

To identify regions with chromatin accessibility specifically associated with one topic compared to all other topics, we conducted topic-level differential accessibility analysis using the “grade of membership” (GoM) method implemented in the de_analysis function. The log-fold change relative to the null was calculated without applying adaptive shrinkage. For each topic, we selected the top 30,000 peaks based on two-tailed p-values (p-values were below 0.05 in all cases) and then filtered for positive log-fold changes, leading to peak sets containing slightly fewer than 30,000 regions. The peak sets were defined as topic specific peaks and used in downstream analysis including sLDSC. We validated that all topic-specific peaks met the p-value cutoff of 0.05 for each topic.

For each topic, gene scores were calculated by aggregating topic loadings (z-score obtained from GoM analysis) for each accessible region/peak (weighted by bi-directional exponential decay from the gene TSS). The gene scores were subsequently normalized by the L2 norm of the weights. Gene set categories (H: hallmark gene sets, C2: curated gene sets, C5: ontology gene sets) from the Molecular Signatures Database (MSigDB) were retrieved and analyzed for each topic-specific gene score vector using the msigdbr package (v7.5.1)^91^. Gene set enrichment analysis was performed using the GSEA function in the clusterProfiler package^92^.

### Transcription factor motif enrichment

We followed the ArchR pipeline to perform enrichment analysis of TF binding motifs within peak sets. We annotate each peak within the union peak set with the position weight matrices (PWMs) from the CisBP^93^ database using the addMotifAnnotations function. Background peaks were computed, controlling for total accessibility and GC content, via the addBgdPeaks function, and per-cell TF ChromVAR activity scores were calculated using the addDeviationsMatrix function. To identify positive TF regulators, we aggregated cells based on cell type, tissue location, and inflammation condition. We then correlated the integrated gene score matrix (scATAC gene score integrated with scRNA data) with the TF activity score matrix across these aggregates. TFs meeting the following criteria were identified as positive regulators: correlation > 0.4, inter-cluster deviation z-score difference > 0.25, and adjusted p-value < 0.01. We further assessed the enrichment of TF motifs in specific peak sets (cell type-specific, inflammation-specific, or topic-specific) using the hypergeometric test, comparing motif overlap with peak sets against random expectations. TFs were prioritized by filtering for positive regulators (Supplementary Table 3).

## Genotyping and quality control

ATAC-seq generates appreciable genome coverage (2.8-24) and we used GLIMPSE2^94^ to determine the genotypes of individuals. The b38 reference panel from the 1000 Genomes Project^95^ was downloaded from the EBI 1000 Genomes FTP site and filtered to retain only SNPs, removing multiallelic records. To improve processing efficiency, genotype data was excluded from the reference VCF files. The GLIMPSE2_chunk function was then employed to define chunks for imputation and phasing. A binary reference panel was constructed using the filtered reference panel, along with the corresponding genetic map and imputation regions from the previous step. With sorted BAM files from the Cell Ranger pipeline as input, the GLIMPSE2_phase function was applied to impute genotypes within each defined region using the constructed reference panel. Finally, the imputed chunks were ligated using GLIMPSE2_ligate tool, and Eagle (v2.4.1) was subsequently used for further phasing, utilizing the same reference panel^96^. To impute individual-level genotypes for subjects with multiple samples, the sample-specific BAM files were merged into individual-specific BAM files using the samtools merge function. Both sample-level and individual-level genotypes were generated to validate the robustness of the pipeline. The quality of individual-level BAM files was assessed by evaluating coverage depth (Extended Data Fig. 6a), calculated using samtools depth, and effective coverage (Extended Data Fig. 6b), as described in Li et al.^97^, to ensure high imputation accuracy. Non-reference concordance in chromosome 1 was examined in Extended Data Fig. 6c, with all pairwise comparisons of samples from the same individual achieving concordance scores exceeding 0.9, further validating the accuracy of the imputation.

### caQTL calling

We used the RASQUAL^98^(v1.1) to compute caQTLs in each cell type. For each individual ID and cell type, we aggregated the scATAC data into pseudo-bulk samples. Only pseudo bulk samples containing more than 20 cells were retained, yielding 19 cell types for analysis. We extracted insertion count matrices for each pseudo bulk sample based on the union peak set across all cell types from ArchR analysis. Allele specific counts were extracted from individual-level bam files and vcf files, with the VCFs filtered to include only variants with a minor allele frequency of at least 5% in our samples, using the createASVCF script. The rasqualTools pipeline (https://github.com/kauralasoo/rasqual/tree/master/rasqualTools) was employed to prepare format-compatible count matrices and GC bias-corrected size factors. Covariates were generated using the size-adjusted peak count matrix via the rasqualMakeCovariates function. We also included age, sex, and top four ancestry principal components (PCs) derived from genotype data from our samples and the 1000 Genomes Project as additional co-variants.

For each peak in the scATAC-seq union peak set, we tested associations between chromatin accessibility and genotype of all variants within a ±10 kb window. Variant-level p-values were transformed into q-values to account for the number of variants tested per peak. To correct for multiple testing genome-wide, we performed three additional association tests using permuted genotypes and calculated an empirical false discovery rate (FDR) for lead variants by comparing the q-values of the real and permuted association results. We obtained QTL mapping results for all tested SNPs and QTLs with an FDR of 10% or below were deemed significant. To visualize chromatin accessibility across different genotypes for significant QTLs, we performed variance-stabilized normalization on the read counts using the varianceStabilizingTransformation function in DESeq2^99^ and plotted the resulting data as boxplots.

### Variant annotation and TF motif disruption analysis

The variants were annotated with their nearest genes using the matchGenes function implemented in bumphunter (v1.36.0) package^100,101^, based on hg38 genome assembly, with a promoter distance cutoff of 2000 bp. Motif disruption analysis was conducted using the information content method implemented in motifbreakR^102^ (v2.8.0). SNPs exhibiting a TFMPvalue of less than 1e-4 were classified as significant motif-disrupting variants. The motif disruption score was calculated by subtracting the motif binding score of the reference allele from that of the alternative allele.

### mashR

We used mashr^47^ (v0.2.79) to estimate the sharing of caQTLs across different cell types. The effect size (effect size = Pi - 0.5) and standard error (SE = abs(beta/sqrt(Chi-square))) for each SNP was calculated obtained from RASQUAL results, In the mash model fitting, we included all SNPs that were tested across all cell types, with no ’NA’ and infinite effect sizes and standard errors. The QTLs that were significantly called in at least one cell type (FDR<10%) were included in the strong test subset, and 2 million peak-SNP pairs from the remaining set were randomly selected as a control subset. During the query dataset construction, the SNP with the lowest q-value for each peak across cell types was selected. For query SNPs, some were not properly tested in certain cell types because of power issues, thus, missing effect size and standard error values (due to absent insertion or allele quantification) were set to 0 and 1e-6, respectively, similar to GTEx study^103^. The fitted mash model was used to reassess effect sizes and significance of the query SNPs. Finally, the SNPs in each cell type were filtered using a local false sign rate (LFSR) cutoff of 0.05 as significant QTLs.

### caQTL effect size and motif disruption score correlation

For each transcription factor (TF) within each cell type, we first identified the caQTLs that were significant in the respective cell type and exhibited a significant motif disruption effect for the corresponding TF (p-value < 1e-4). We then computed the Spearman correlation between the motif disruption scores and the QTL effect sizes for each TF-cell-type pair. We only included TF motifs with at least five significant motif-disrupting QTLs in a given cell type. The p-values from these correlations were corrected for multiple testing using the Benjamini-Hochberg procedure. For visualization, the coefficients of non-significant correlations (adjusted p-value cutoff = 0.05) were not displayed. This entire workflow was repeated using QTLs identified as significant by the mash method.

### Pairwise caQTL effect size correlation

For each pair of cell types, we selected caQTLs that were significant in either cell type evaluated by mash and retrieved the corresponding effect sizes calculated using the RASQUAL workflow. Missing effect sizes were set to 0. We then computed the Spearman correlation of QTL effect sizes between the two cell types. The correlation p-values were adjusted for multiple testing using the Benjamini-Hochberg procedure. For visualization, the coefficients of non-significant correlations were set to zero (adjusted p-value cutoff = 0.05)

### Heritability enrichment

We used the 1000G European panel^95^ as reference SNPs and HapMap3^104^ SNPs as the regression set, excluding SNPs located in the major histocompatibility complex region. Since these references and GWAS data were available in hg19, we lifted over the union set of chromatin accessibility peaks from hg38 to hg19 using the UCSC LiftOver tool. Peak regions were then extended by 1,000 bp both upstream and downstream for SNP annotation, accounting for biases between the reference panel and scATAC-seq-based peak calling. To assess the enrichment of disease or trait heritability within a specified peak set, we used stratified linkage disequilibrium score regression (sLDSC) with the baseline-LD (v2.2) model^50,67^. As a conservative step to ensure cell-type specificity, we additionally conditioned the analysis on all cell-type-specific peaks^68^. We followed the post-processing pipeline described in Samuel et al.^68^ to calculate the p-value of ρ^∗^ to evaluate the significance of heritability enrichment. ρ^∗^ represents the standardized effect size, indicating the proportionate change in per-SNP heritability associated with a one standard deviation increase in the annotation value. The preprocessed reference files, baseline model and GWAS summary statistics for sLDSC analysis are available from Alkes group: https://console.cloud.google.com/storage/browser/broad-alkesgroup-public-requester-pays/LDSCORE.

### Statistical fine-mapping

The GWAS summary statistics (hg19) for Crohn’s disease provided by de Lange et al.^14^ were utilized for this analysis. Since the sample genotypes and in-sample LD matrices from the meta-analysis GWAS study are not publicly available, we selected two reference panels with matched ancestries to estimate the LD matrix: the 1000G European panel^95^ and the UK Biobank panel^58,59^, as computed in Zhao et al^105^. Fine-mapping was conducted separately using each reference panel. The GWAS summary statistics were preprocessed and harmonized with the LD reference panel using the pipeline in mapgen (v0.5.9)^26^. The genome was partitioned into LD blocks with LDetect^106^, and loci containing at least one SNP with genome-wide significance (p-value < 5e−8) were selected for fine-mapping. We conducted fine-mapping with summary statistics using the sum of single effects (SuSiE) method^107,108^, implemented in susieR (v0.12.35), with a uniform prior and permitting up to ten causal variants per locus. SNPs that mismatched the reference panel were filtered. Loci overlapping the region from 25 Mb to 35 Mb on chromosome 6, corresponding to the HLA region, were identified and excluded from prioritization. We provided the complete fine-mapping results, calculated using the 1000G reference panel, in Supplementary Table 5. Peak-overlapping SNPs with posterior inclusion probabilities (PIPs) greater than 0.2, as determined using both the 1000G and UKB panels, were prioritized and listed in Table 1.

### Enrichment of causal signals in cCREs

The fold enrichment of causal signals in cCREs was calculated using the following formula: (PIP_cCRE/genome_length_cCRE) / (PIP_non_cCRE/genome_length_non_cCRE). Regions in the black list of hg38^34^ were removed in both cCRE regions and non cCRE regions during analysis. To test the significance of the enrichment, we rounded the total PIP in each of the two groups to an integer as causal signals, and compared the number of causal signals among the total number of finemapped variants using Fisher’s exact test.

### Functional annotation and prioritization of fine-mapped SNPs

We annotated fine-mapped SNPs with the cell-type-resolved chromatin accessibility by directly overlapping SNPs with the union set of peaks.

We linked cCREs containing SNPs to their putative target genes using 3 different methods, co-accessibility, ABC-Max method, and nearest TSS. For co-accessibility, we identified co-activity between gene expression and peak accessibility using the addPeak2GeneLinks function, selecting peak-gene links with a correlation score greater than 0.5, as provided in Supplementary Table 2. Most Fine-mapped SNPs overlapping with cCREs were annotated with the co-active gene. Additionally, we annotated fine-mapped SNPs using the ABC-Max method. We included the peak-gene links from biosamples that have ABC enhancers enriched for IBD fine-mapped variants, as well as the “small_intestine_fetal-Roadmap” biosample provided in Nasser et al^70,71^. We extended the ABC-Max method to annotate QTL caPeaks by evaluating the ABC scores of all ABC enhancers overlapping the caPeak, assigning the caPeak to the target gene with the highest ABC score. Finally, in cases where neither method provided clear target genes or assignments were ambiguous, we provided the gene with the closest TSS as the likely target. For the small subset of regulatory candidate variants, we also manually assessed the gene assignment (Supplementary Table 2).

To identify GWAS variants that are caQTLs in our dataset, we first calculated LD between variants using PLINK (v1.9)^109^ based on a merged panel combining our samples and 1000G European samples. Variants exhibiting strong pairwise LD (r² > 0.8) were classified as overlapping. QTL SNPs that overlapped with fine-mapped SNPs were provided in Supplementary Table 6.

---

**Extended Data Fig. 1.**
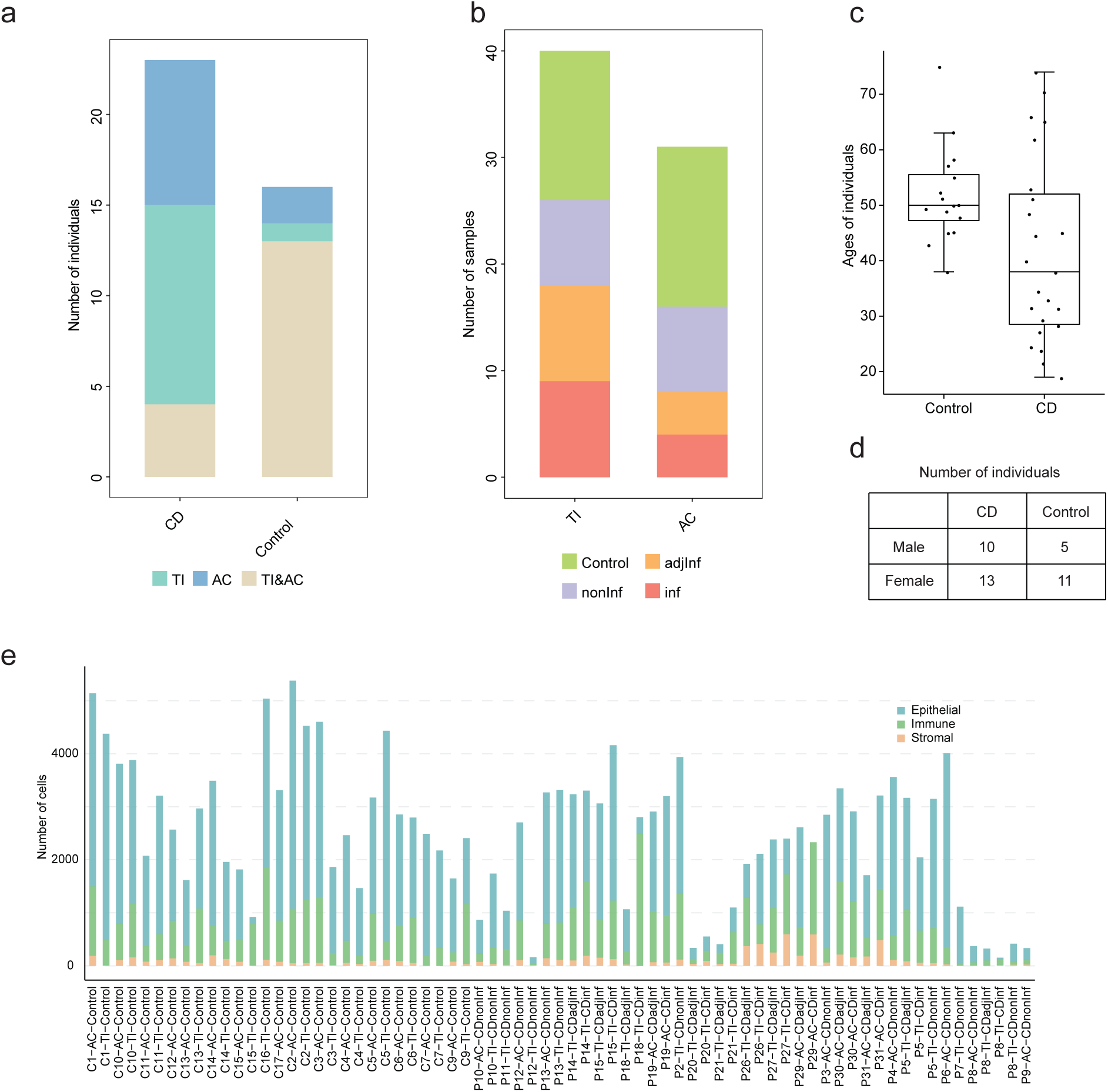
Sample overview. **a**, The number of individuals that have samples collected from only terminal ileum (TI), only ascending colon, or both regions (TI & AC). **b**, The number of samples in different disease conditions. Control: samples from the control group. nonInf: samples without active inflammation from CD patients. adjInf: non-inflamed tissue adjacent to inflamed site. inf: inflamed tissue. **c**, Distribution of ages of individuals. **d**, Number of individuals stratified by sex and disease condition. **e**, The number of cells across different lineages in each sample.

**Extended Data Fig. 2.**
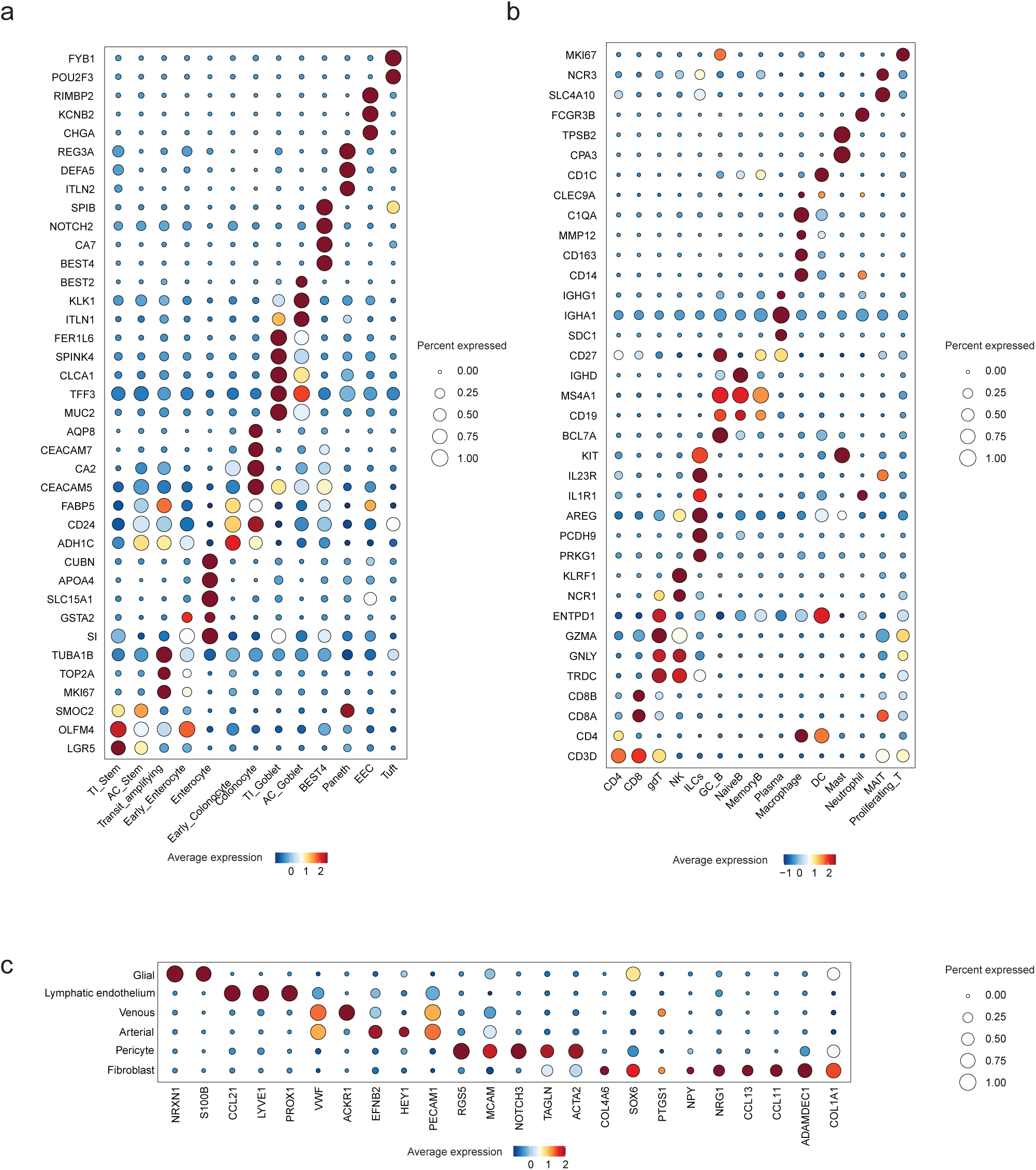
Markers for scRNA-seq data annotation. **a**, **b**, **c**, Expression of markers for different cell types in each lineage (a, epithelial. b, immune. c, stromal.). The color shows scaled mean expression in each cell type. The dot size represents the proportion of cells expressing that marker.

**Extended Data Fig. 3.**
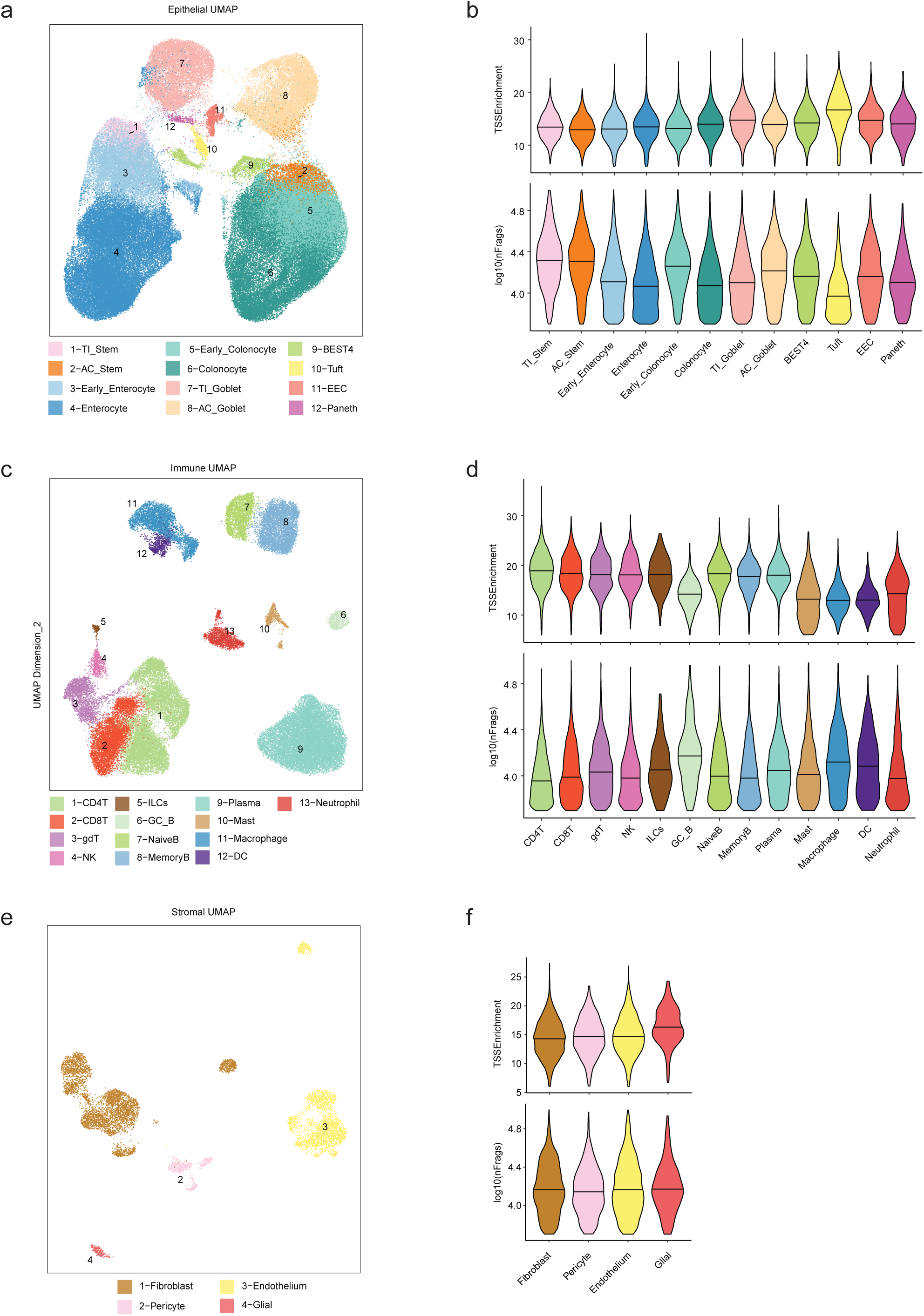
Cell type annotation and quality control in scATAC-seq. Visualization of different cell types in lineage-specific UMAP embedding and the distribution of single cell TSS enrichment scores and fragment counts in each cell type (a, b, epithelial. c, d, immune. e, f, stromal.).

**Extended Data Fig. 4.**
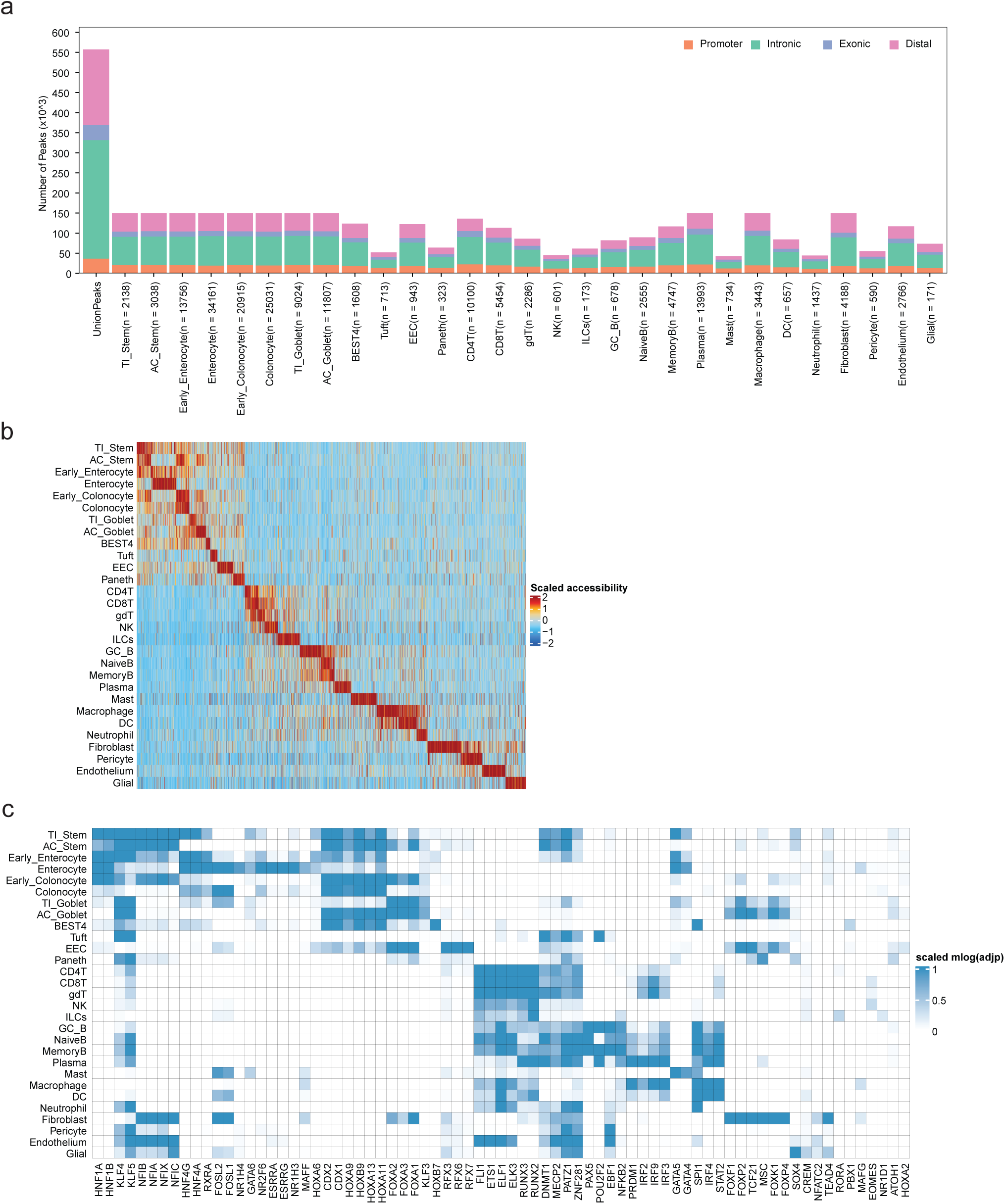
Cell type specific DARs and TFs. **a**, The number of peaks in the union peak set and the peakset called using MACS2 for each cell type. **b**, Cell-type-specific differentially accessible regions (DARs). Each column is a DAR. **c**, Cell-type-specific transcription factor motifs enriched from cell-type-specific DARs. The -log10(adjusted p-value) for motif enrichment was capped at 100, with any values exceeding 100 being set to 100, and scaled by the maximum value per TF (right).

**Extended Data Fig. 5.**
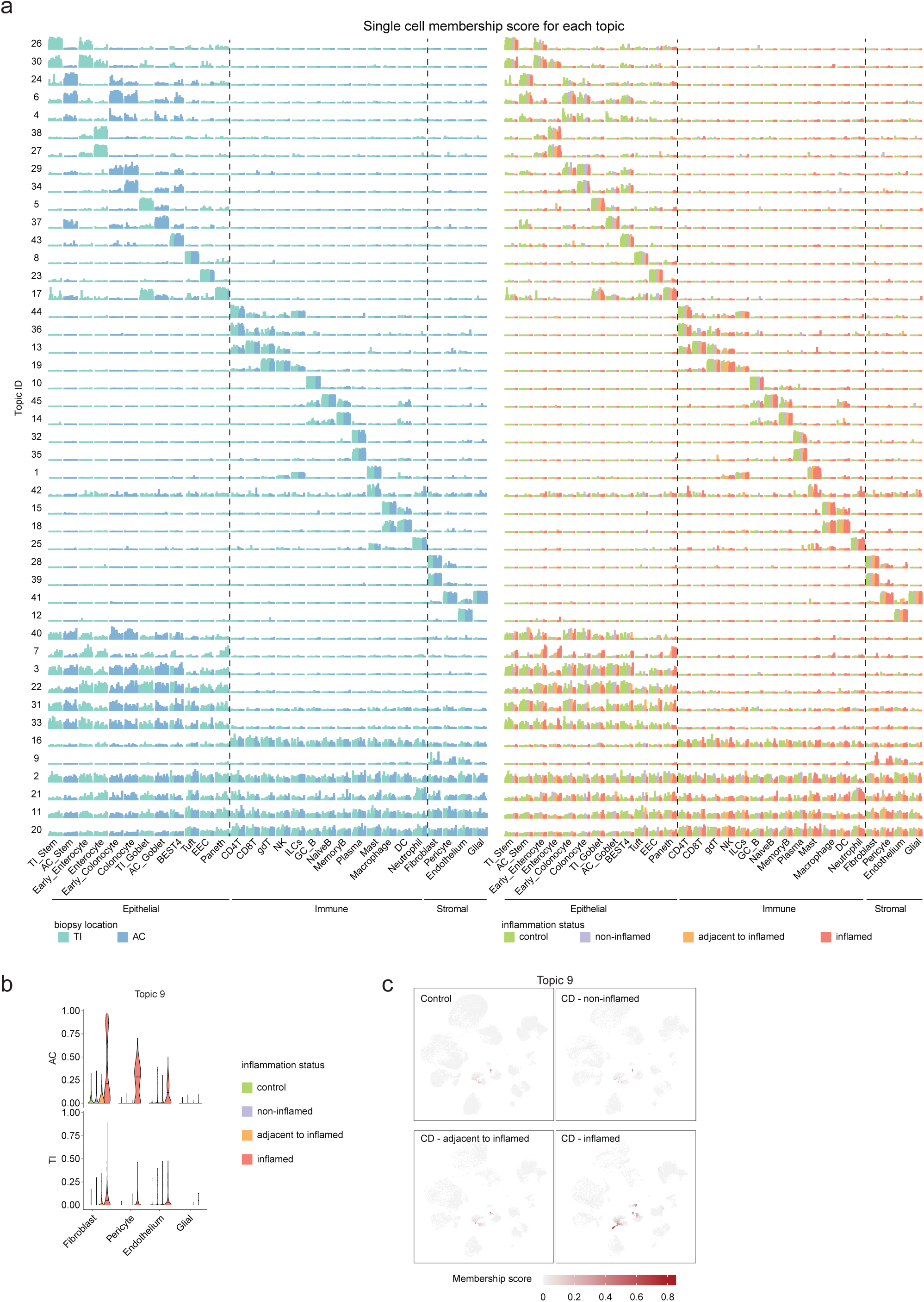
Structure plots of all topics. a, Structure plots displaying proportion of topics in individual cells. The cells are colored by biopsy location (left) or biopsy inflammation status (right). Each column represents a single cell and cells are grouped by cell types. Topic scores are scaled by the maximum value per row. **b**, Proportion of topic 9 membership for each cell. Cells grouped by cell type, tissue location, and inflammation status. **c**, UMAP plots displaying proportion of topic 9 membership for each cell, split by inflammation status.

**Extended Data Fig. 6.**
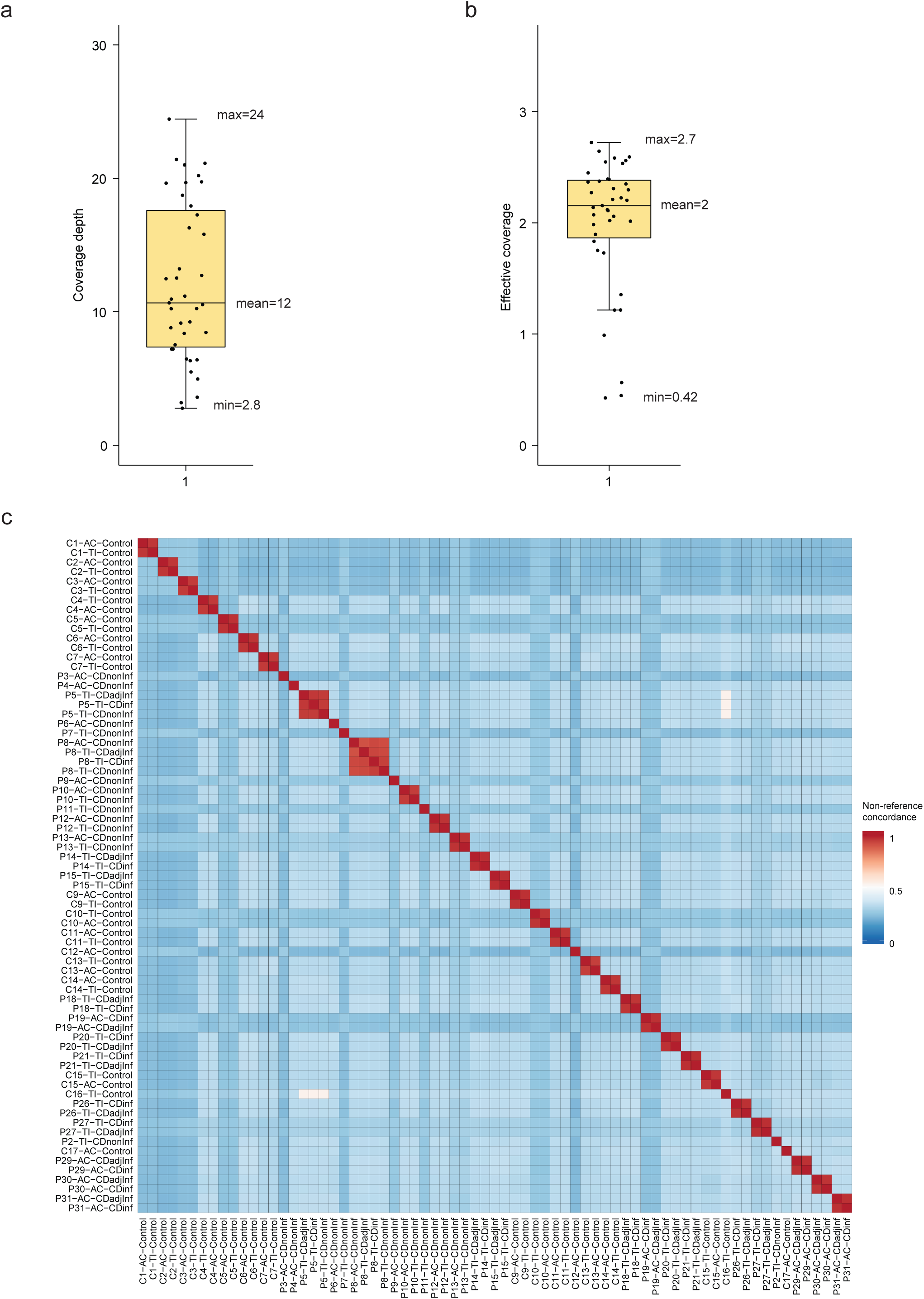
Quality control of genotyping. a, Distribution of coverage depths of individual-level BAM files. **b,** Distribution of effective coverages of individual-level BAM files. **c**, Non-reference concordance of imputed variants in chromosome 1 between each pair of samples.

**Extended Data Fig. 7.**
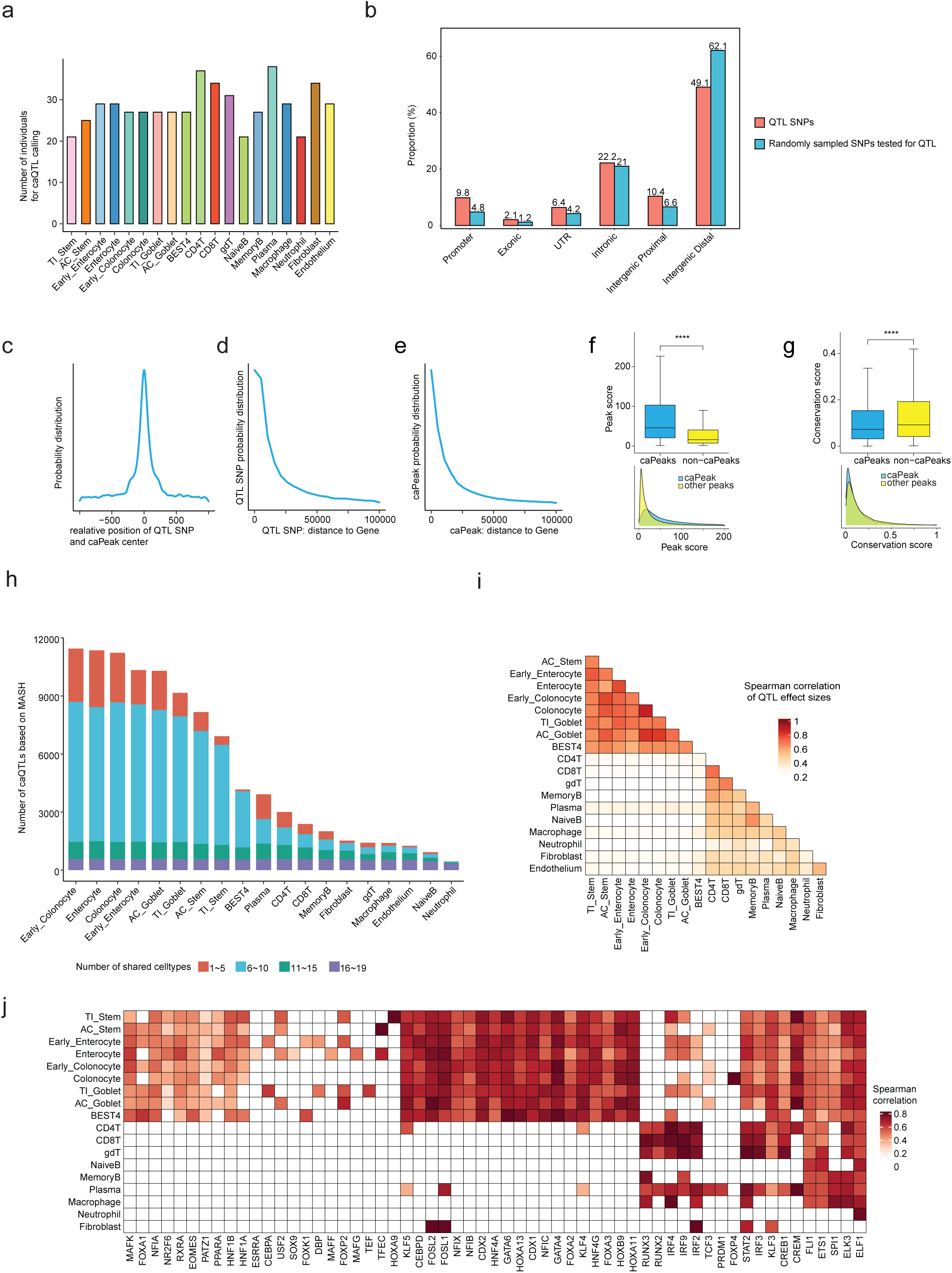
Evaluation of caQTLs. **a**, Number of individuals used for caQTL calling in each cell type. **b**, Proportion of significant QTL SNPs and randomly sampled SNPs across genomics annotations. UTR: Untranslated Region, Promoter: within 2 kb of transcription start site of the nearest gene, Intergenic proximal: within 5 kb of the 5’ end of the nearest gene. Intergenic distal: more than 5 kb away from the 5’ end of the nearest gene. **c**, Distribution of the distances between each QTL SNPs and associated caPeaks. **d**, Distribution of the distances between each QTL SNPs and the nearest gene. **e**, Distribution of the distance between each caPeak and the nearest gene. **f**, Peak scores obtained from MACS2 peaking calling for caPeaks and non-caPeaks. **g**, Sequence conservation (PhastCons score) for caPeaks and non-caPeaks. **h**, Number of caQTLs significant in each cell type assessed by mashR (LFSR<0.05). The number of shared cell types for caPeaks are indicated by color. **i**, Spearman correlation coefficients of QTL effect sizes for all pairwise cell type comparisons based on mash results. **j**, Heatmap of Spearman correlation coefficients between QTL motif breaking scores and effect sizes for each TF-cell type pair. Coefficients are only displayed for significant correlations (FDR <0.05). The QTLs for this analysis were determined to be significant by mashR.

**Extended Data Fig. 8.**
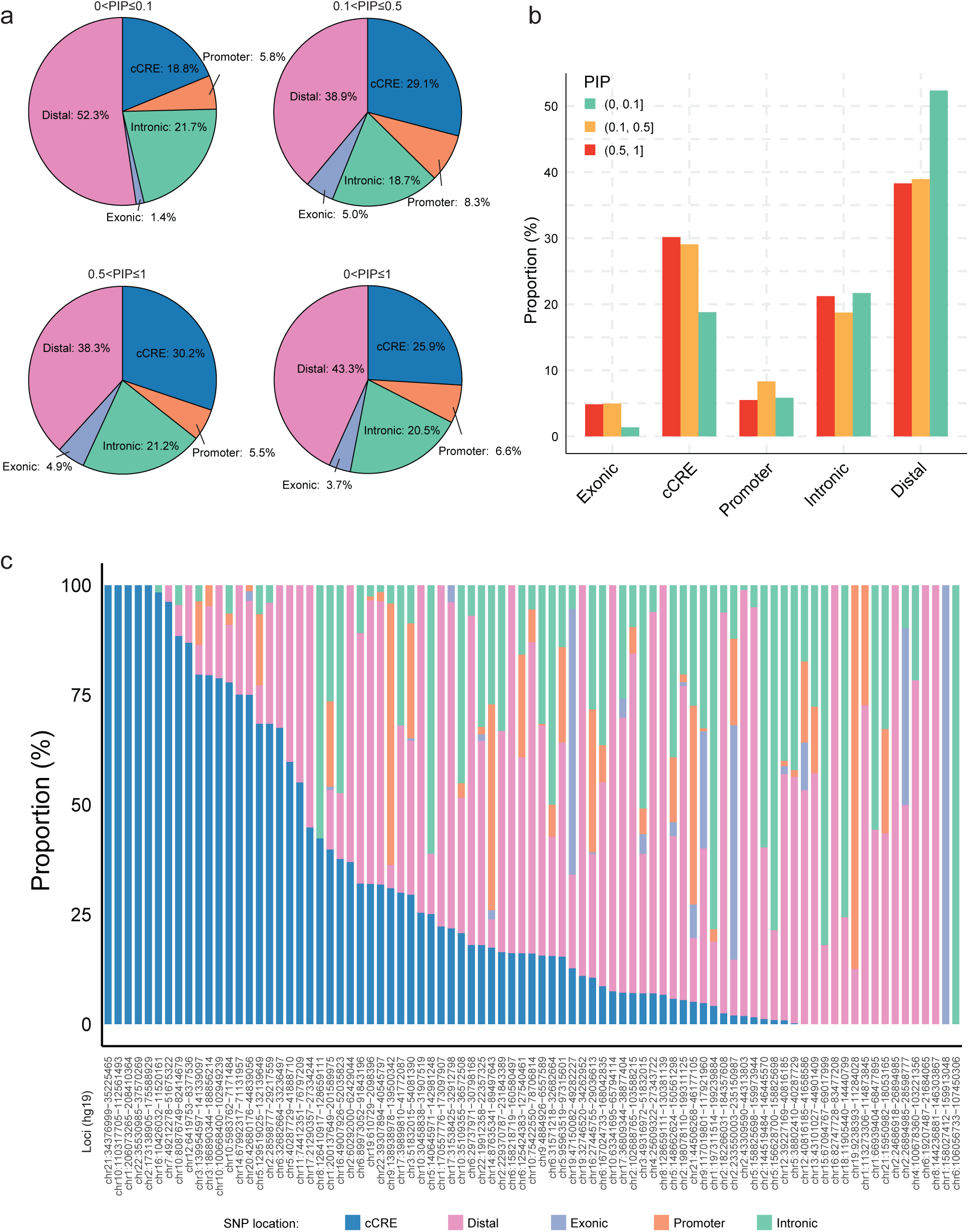
Overlap of fine-mapped SNPs with genomic regions. a, **b** Proportion of summed PIPs in each type of genomic region. SNPs within different ranges of PIP values were calculated separately. **c**, Proportion of summed PIPs in different types of genomic regions at each individual locus.

